# The immune response to SARS-CoV-2 as a recall response susceptible to immune imprinting: a prospective cohort study

**DOI:** 10.1101/2024.08.11.24311358

**Authors:** Daniel Alvarez-Sierra, Mónica Martínez-Gallo, Adrián Sánchez-Montalvá, Marco Fernández-Sanmartín, Roger Colobran, Juan Espinosa-Pereiro, Elísabet Poyatos-Canton, Coral Zurera-Egea, Alex Sánchez-Pla, Concepción Violan, Rafael Parra, Hammad Alzayat, Ana Vivancos, Francisco Morandeira-Rego, Blanca Urban-Vargas, Eva Martínez-Cáceres, Manuel Hernández-González, Jordi Bas-Minguet, Peter D. Katsikis, Aina Teniente-Serra, Ricardo Pujol-Borrell

## Abstract

**Background:** The antibody response to SARS-CoV-2 does not follow the immunoglobulin isotype pattern expected in a primary response and is inconsistent with the current interpretation of COVID-19 immunopathology as the result of a primary infection. To better understand the immune response to SARS-CoV-2, it is essential to determine whether it is primary or secondary (or recall). The analysis of highly granular immunological variable trajectories of a homogeneous cohort of patients receiving standardised medical care should discern between primary and secondary responses.

**Methods:** This is a prospective cohort study of 191 SARS-CoV-2 infection cases and 44 healthy controls from the second wave of COVID-19 in the Barcelona area. The study stratified patients by severity and analysed the trajectories of SARS-CoV-2 antibodies and multiple immune variables for features associated with primary and recall immune responses.

**Findings:** Isotype-specific antibody trajectories to SARS-CoV-2 proteins revealed a pattern of recall response in 94·2% of cases. In these cases, the detailed trajectories of plasmablasts, B cells, cTfh high-resolution subsets, and cytokines were consistent with a secondary response. The transcriptomic data indicated that this cohort is strictly comparable to contemporary cohorts.

**Conclusions:** In most cases, the immune response to SARS-CoV-2 is a recall response. This opens the possibility that most COVID-19 cases are subjected to immune imprinting by endemic coronavirus, which, in turn, can contribute to severity by interfering with the immune response to SARS-CoV-2 and by antibody-dependent enhancement. Considering the immune responses to SARS-CoV-2 secondary provides a better perspective to interpret COVID-19 pathology.

**Funding:** Grants COV20/00416, Cov20/00654, and COV20/00388 from Instituto de Salud Carlos III (ISCIII), Madrid, Spain, co-financed by the European Regional Development Fund (ERDF).

## INTRODUCTION

The COVID-19 pandemic has resulted in over 775 million cases of clinical infections and 7 million deaths worldwide as of October 2024. (https://data.who.int/dashboards/covid19). Even after unprecedented extensive research, some critical questions still demand answers. One of the most debated issues is what causes tissue damage in severe COVID-19. Such damage could be due to virus cytopathic effects, immune-mediated pathology, or a combination.^1^ Understanding the pathogenesis of acute COVID-19 will also help to advance research on long COVID, where uncertainty remains on whether symptoms are due to persistent infection, a dysregulated immune response, or other still-to-be-defined mechanisms. ^2^

The clinical severity of COVID-19 varies greatly, ranging from no symptoms to death. A proportion of this variability can be attributed to some well-established host risk factors identified in the early stages of the pandemic, in order of importance: age, sex, comorbidities such as hypertension, diabetes, obesity, chronic lung disease, chronic kidney disease, and immunosuppression. ^3^ Mutations targeting the type I IFN pathway have been identified, accounting for approximately 3% of severe cases.^4,5^ Autoantibodies to type-I IFNs explain a proportion of severe cases.^6^ Collectively, dysfunction of the Type I IFN pathway by a gene defect or autoantibodies may explain up to 20% of severe cases. ^5^ A recent COVID-19 Host Genetics Initiative (HGI) meta-analysis identified 21 loci associated with susceptibility and 40 loci associated with COVID-19 hospitalisation. These loci accounted for 1·2% and 5·8% of phenotypic variation, respectively.^7^ The total heritability of severity has been estimated to be 41% (9), but, at present, a significant proportion of COVID-19 severity attributable to the host remains unexplained, as revealed in many multivariate analyses, including ours. ^8^

Some early reports indicated that in COVID-19, the response of IgA and IgG antibodies to SARS-CoV-2 proteins preceded or was contemporary to IgM.^9,10^ This is surprising because, in a typical primary immune response, IgM always precedes IgG and IgA.^11,12^ One possible explanation is the interference of the immune response to SARS-CoV-2 proteins by pre-existing cross-reactive immunity to endemic coronavirus. ^13^

Even if there is solid evidence that many subjects not previously exposed to SARS-CoV-2 have immunity to SARS-CoV-2 proteins.^14–17^, These observations lead to postulate that the immune response to SARS-CoV-2 was interfered with by pre-existing immune response by a mechanism known as Immune Imprinting (IP), original antigenic sin, or antigenic addiction, terms that are not precisely synonymous (see discussion). ^18^ Limited but robust retrospective studies indicate that IP plays a crucial role in COVID-19.^13,19–21^ Still, neither IP nor antigenic original sin has been fully incorporated in the prevailing models of COVID-19 immunopathology. ^1,22,23^ Demonstrating IP requires comparing pre- and post-COVID-19 paired serology samples and showing that the infection evokes a response to endemic Common Cold Coronavirus (CCCV) rather than to SARS-CoV-2 which is challenging due to the limited availability of paired samples and epidemiological data. Interpreting the results from these studies is also hindered by the potential overlap of cross-priming, IP, and de novo priming.

In the course of a comprehensive investigation of the immune response to SARS-CoV-2 in a prospective cohort of unvaccinated COVID-19 patients from the Barcelona metropolitan area second wave, we found evidence that in almost 95% of patients, the serological and cell subset features fit those of recall immune response. These features were notably more prominent in moderate and severe cases than in asymptomatic and mild cases.

## METHODS

### Patients

The cohort of patients participating in the study was recruited by the attending physicians of the participating hospitals (Hospital Universitari Bellvitge (HUB), Hospital Universitari Germans Trias i Pujol, (HUGTP); Hospital Universitari Vall d’Hebron (HUVH) upon admission or at primary care centres during the first contact. In the latter scenario, health workers visited patients’ homes to collect clinical and blood samples. Detected asymptomatic household sharing cases were recruited. Controls were recruited among the Catalonian Blood and Tissue Bank (www.bst.cat) blood donors who had a more similar age and sex composition as the COVID-19 patients. All participants were informed of the project’s objectives and signed the consent forms. Blood samples were collected into citrate, EDTA, sera separation, and Tempus® tubes (Becton-Dickinson Inc, NJ, USA).

A Research Electronic Data Capture (REDCap) database^24^ was generated to collect clinical and laboratory data. Information was introduced in 182 fields, including demographics, medical treatment history, comorbidities, Charlson’s and SOFA indexes, initial symptoms, vital signs, physical examination, follow-up including oxygen requirements and therapy, and clinical chemistry data. The clinical data and blood samples were collected at the beginning and at two more time points. For all patients, the date of symptom onset was recorded and used to calculate “days from symptom onset” (DFSO) for each observation. The length of hospital stays (LOS), ICU stay, oxygen supplementation, and ventilation support were also recorded.

Clinical severity categories were determined by the highest score during the follow-up period using the World Health Organization (WHO) 8-point COVID-19 disease clinical progression score.^25^ The scores correspond to phenotypic categories: 0 no clinical or virological evidence of infection, 1: no limitation of activities, 2: limitation of activities, not requiring hospitalisation, 3: hospitalised without oxygen requirement, 4: oxygen administered via a mask or nasal prongs, 5: non-invasive ventilation or high-flow oxygen, 6: intubation and mechanical ventilation, 7: ventilation and additional organ support and 8: deceased. In most analyses, patient classification was simplified as asymptomatic (score 1), mild (score 2), moderate (score 3-4), and severe (score of 5 to 8). Moderate and severe patients were all hospitalised.

### Clinical Laboratory Tests

SARS-CoV-2 was detected by a real-time multiplex RT-PCR assay (Laplet 2019-nCoV Assay, Seegene, South Korea) in samples from nasal or pharyngeal swabs. Microbiological and clinical chemistry samples were processed by automatic analysers integrated into continuous lines with automatic cold storage that ensured sample integrity.

### Immunological Tests

#### Cytokines and related proteins

The levels of CCL2, CXCL10, GM-CSF, IFN-α, IFN-γ, IL-2, IL-4, IL-6, IL-7, IL-10, IL-12p70, IL-13, IL-15, IL-17A, TGF-β1, TNF-α, granzyme-B and IL-1RA were measured in sera using the ELLA microfluidic platform (Biotechne®, Minneapolis, MN, USA). Calprotectin was measured by CLIA (Quantaflash®, Werfen, Barcelona, Spain).

#### SARS-CoV-2 serology

Antibodies of the three immunoglobulin isotypes IgM, IgG, and IgA against SARS-CoV-2 main protease (Mpro), nucleocapsid (NP), Spike (S) protein, and the RBD portion of the Spike protein were measured in serum samples using a commercial kit (SARS-COV-2 MULTIPLEX®, IMMUNOSTEP, Salamanca, Spain).^26^ In some analyses, the sum of antibody levels for each isotype to Mpro, NP, and S proteins were used to score each individual’s overall isotype-restricted response to SARS-CoV-2, annotated as SARS-CoV-2 IgA, SARS-CoV-2 IgG, and SARS-CoV-2 IgM.

#### Type 1 IFN autoantibody measurement

The ELISA was performed as previously described.^6^ In brief, ELISA plates were coated overnight at 4°C with 1 μg/mL of recombinant human interferon-α (rhIFN-α, Miltenyi Biotec, Germany) or rhIFN-ω. After washing, the plates were blocked with PBS 1X supplemented with 5% skim milk powder, and then 1:50 dilutions of serum samples from the patients and controls were added and incubated for 2 hours at RT. After washing, specific Fc goat anti-human IgG was added. Finally, SureBlue™ TMB 1-Component Microwell Peroxidase Substrate (Biogen, USA) was added, and the optical density was measured using a Varioskan LUX Multimode Microplate Reader (ThermoFisher Scientific, USA) at 450/630 nm. Positive controls and blanks were used as internal quality controls.

#### High-dimensional flow cytometry

Whole blood was processed as detailed in the article by Fernandez-Sanmartin et al. ^27^ Red blood cells were lysed, and freshly obtained cells were stained with a 36-color antibody panel (Supplemental Table 1) and analysed using a 5-laser Aurora spectral flow cytometer (Cytek Biosciences). ^27^ Unsupervised statistical inferences of the data were computed by OMIQ data analysis software (www.omiq.ai), UMAP was utilised for dimensionality reduction, and flowSOM for clustering (see Results).

#### Transcriptomic profiling by Nanostring

250 ng of total RNA from PBMC, quantified using the NanoDrop 2000 (Thermo Scientific), was directly hybridised (at 65°C for 18 h) with a mixture of biotinylated capture probes and fluorescently labelled reporter probes complementary to target sequences. Gene expression values were first normalised to the positive controls and then normalised according to the nCounter Expression Data Analysis Guide (mAN-C0011-02). The nCounter® Human Immunology Host Response Panel was used for this study (https://nanostring.com/products/ncounter-assays-panels/immunology/host-response).

#### Statistical analysis

Analysis was conducted in the R environment version 4·3·2 and R Studio. The distribution of variables was determined to apply the appropriate type of test. All tests considered two-tailed distributions to calculate the *p*-value, adjusted by Bonferroni except where otherwise stated; *p* values <0·05 were considered significant. In LOESS smoothed regression curves, 95% CI are represented unless otherwise noted. Sex as a biological variable was considered in every statistical analysis but reported separately only when significant differences were detected.cThe analysis was carried out or supervised by the bioinformatic and statistical analysis unit of Vall d’Hebron Research Institute (VHIR) (https://vhir.vallhebron.com/ca/suport-la-recerca/unitat-destadistica-i-bioinformatica-ueb).

### ETHICAL

The institutional ethics board approved this project of the institutions (HUB: 120/20; HUGTP: ImmuneProfile-COVID19 REF.CEI PI-20-218 and HUVH: Protocol number VH, PR(AG)242/2020).

## RESULTS

### Features of the Barcelona COVID second-wave cohort

The final cohort included 191 COVID-19 cases and 44 blood donors as controls recruited in 2020, during the second wave of COVID-19 in Barcelona, before the vaccination campaign started in January 2021.^28^ Of the patients, 38 were asymptomatic, 49 mild, 64 moderate, and 40 severe. The study’s design is depicted in Fig. 1. The inclusion criteria were being above 18 years of age, having a confirmed virological diagnosis of SARS-CoV-2 by PCR test, and agreeing to participate. In the case of healthy blood donors, cases with a history of clinical COVID-19 or a positive serological test in our assay were excluded.

**Figure 1.**
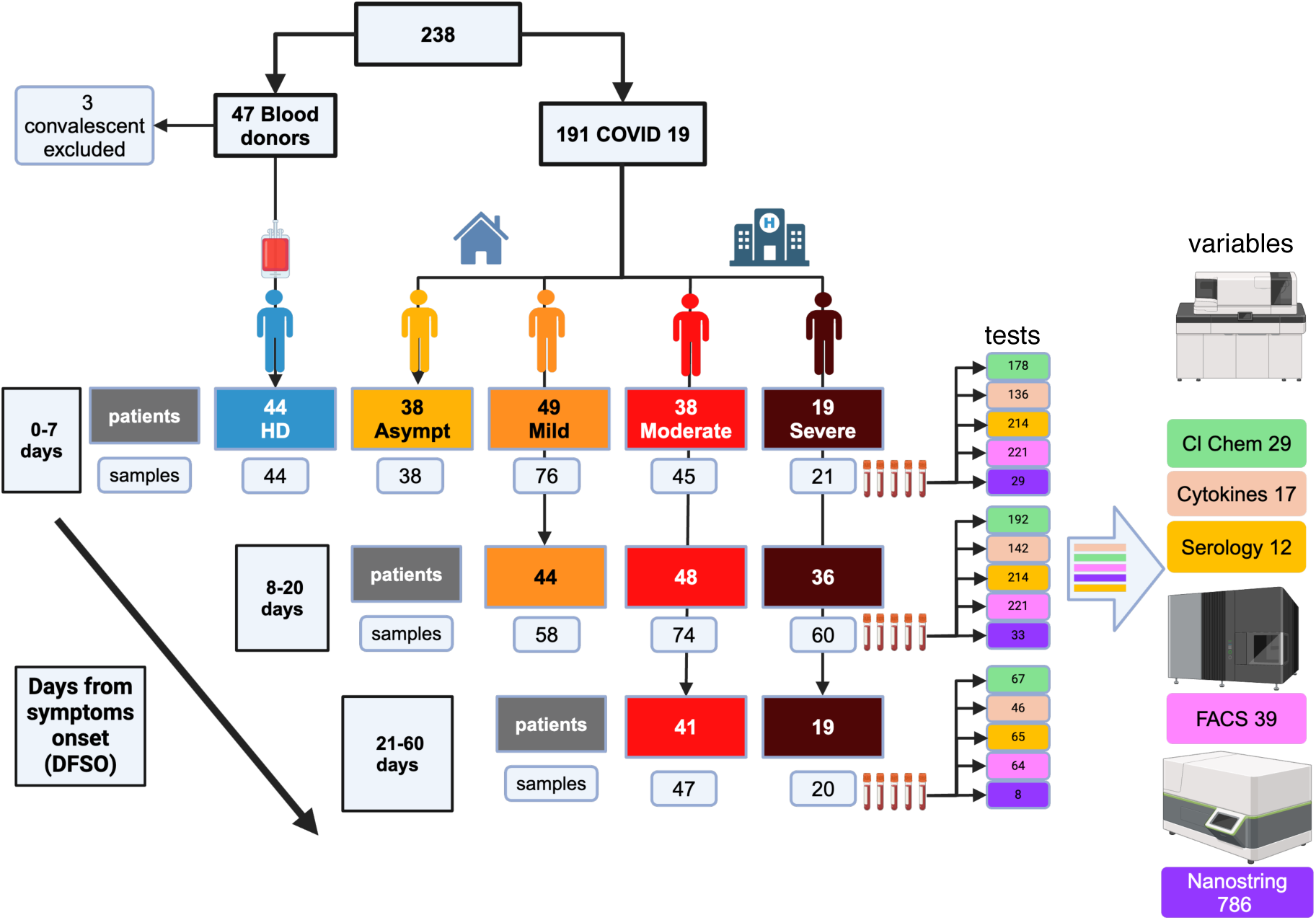
Patients work flow. Study design and patient flow chart. The figure summarises the patient groups, the timeline, and the number of variables measured in each group. Clinical data were available from all 235 individuals, but due to financial limitations, cytokine and Nanostring tests were applied to approximately 67 % and 15 % of the samples, respectively (see text). DFSO, days from symptoms onset; Asympt, asymptomatic cases; see text in section “Features of the Barcelona COVID second-wave cohort” of results for details. Large coloured boxes, patient numbers; small coloured boxes, sample numbers; coloured boxes on the right edge, number of samples by period and type of test; the coloured boxes under the equipment icons give the test colour code.

Table 1 shows the details of cases by period, severity, samples, and tests. For the analysis of results, we have used three time periods based on the days from symptoms onset (DFSO), DFSO1 0-7 days, DFSO2 8-20 days, and DFSO3 21-107 days. The number of patients per period was 101, 128, and 60, and the number of samples per period was 139, 188, and 65, respectively. Of the 483 samples, only 13 (2.7%) were collected at DFSO >60. Due to financial constraints, cytokines were measured in 67 %, and RNA profiles (Nanostring®) in 15% of the selected samples to represent each severity and period group.

**Table 1.** Patients and sample distribution across the follow-up periods.

|  |  | Periods |  |  |  |  |  |  |  |  |  |  |  |  |  |  |  |  |  |
| --- | --- | --- | --- | --- | --- | --- | --- | --- | --- | --- | --- | --- | --- | --- | --- | --- | --- | --- | --- |
|  |  | DFS01 or DFS0_A or DFS0_C (0-7 days) |  |  |  |  |  | DFS02 (8-20 days) |  |  |  |  |  | DFS03 (21-107 days) |  |  |  |  |  |
|  |  | Patients | Samples |  |  |  |  | Patients | Samples |  |  |  |  | Patients | Samples |  |  |  |  |
| Groups, total patients | n | n | Cl Chem | Cytokines | Serology | Flowcyt | Nanostr | n | Cl Chem | Cytokines | Serology | Flowcyt | Nanostr | n | Cl Chem | Cytokines | Serology | Flowcyt | Nanostr |
| Control | 44 | 44 | NA | 24 | 38 | 44 | 6 | NA | NA | NA | NA | NA | NA | NA | NA | NA | NA | NA | NA |
| Asympt | 38 | 38 | 38 | 28 | 37 | 37 | 6 | NA | NA | NA | NA | NA | NA | NA | NA | NA | NA | NA | NA |
| Mild | 49 | 49 | 74 | 47 | 74 | 76 | 10 | 44 | 58 | 41 | 58 | 57 | 10 | ND | ND | ND | ND | ND | ND |
| Moderate | 64 | 38 | 45 | 24 | 45 | 45 | 3 | 48 | 74 | 56 | 73 | 70 | 13 | 41 | 47 | 30 | 46 | 45 | 5 |
| Severe | 40 | 19 | 21 | 13 | 20 | 19 | 4 | 36 | 60 | 45 | 57 | 58 | 10 | 19 | 20 | 16 | 19 | 19 | 3 |
| Totals | 235 | 188 | 178 | 136 | 214 | 221 | 29 | 128 | 192 | 142 | 188 | 185 | 33 | 60 | 67 | 46 | 65 | 64 | 8 |

This table summarises the period of patient enrollment, observation, samples, and variables measured in them. In the top periods, in the bottom table the samples in the order they were taken, A, B and C. Notice that 26 moderate and 21 severe contributed their samples in period DFS02; also that while 107 contributed three samples, 34 two and 14 one. See text section results “Features of the Barcelona COVID second-wave cohort” for details and justification. Cl.Chem = Clinical Chemistry; Flowcyt = Flowcytometry; Nanostr = Nanostring®

Out of the 106 hospitalised patients, 31 (29·2%) required admission to the ICU, and the median length of stay in the hospital (LOSH) was 15 days [7 - 25]. Their ICU stays averaged 14 days [7 - 27]. The demographic, clinical, and laboratory data are in Tables 2 and 3. The overall mortality during the follow-up period was 6/191 (3·1%).

**Table 2.** Patients contribution per period.

| **DFS01** |  |  |  |
| --- | --- | --- | --- |
| Severity | Patients | Samples | Patients that contributed two samples |
| control | 44 | 44 | 0 |
| asymptomatic | 38 | 38 | 0 |
| mild | 49 | 76 | 27 |
| moderate | 38 | 45 | 7 |
| severe | 19 | 21 | 2 |
| Total | 188 | 224 |  |
| **DFS0 2** |  |  |  |
| mild | 44 | 58 | 14 |
| moderate | 48 | 73 | 25 |
| severe | 36 | 58 | 22 |
| Total | 128 | 189 |  |
| **DFS0 3** |  |  |  |
| moderate | 41 | 47 | 6 |
| severe | 19 | 20 | 1 |
| Total | 60 | 67 |  |

This table clarifies the number of patients that contributed more than one sample per a given time period in the trajectory regression analysis.

**Table 3.**
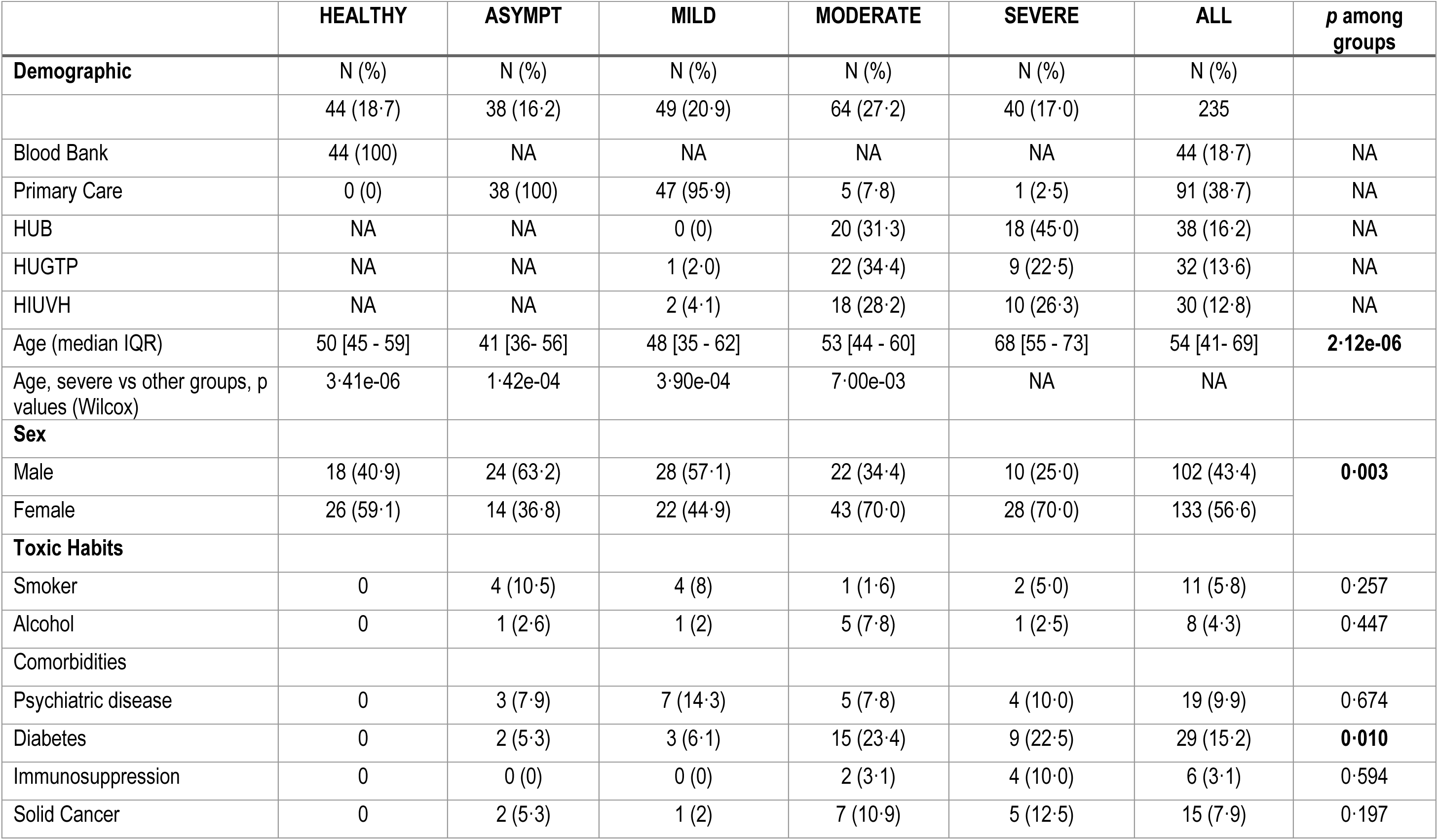
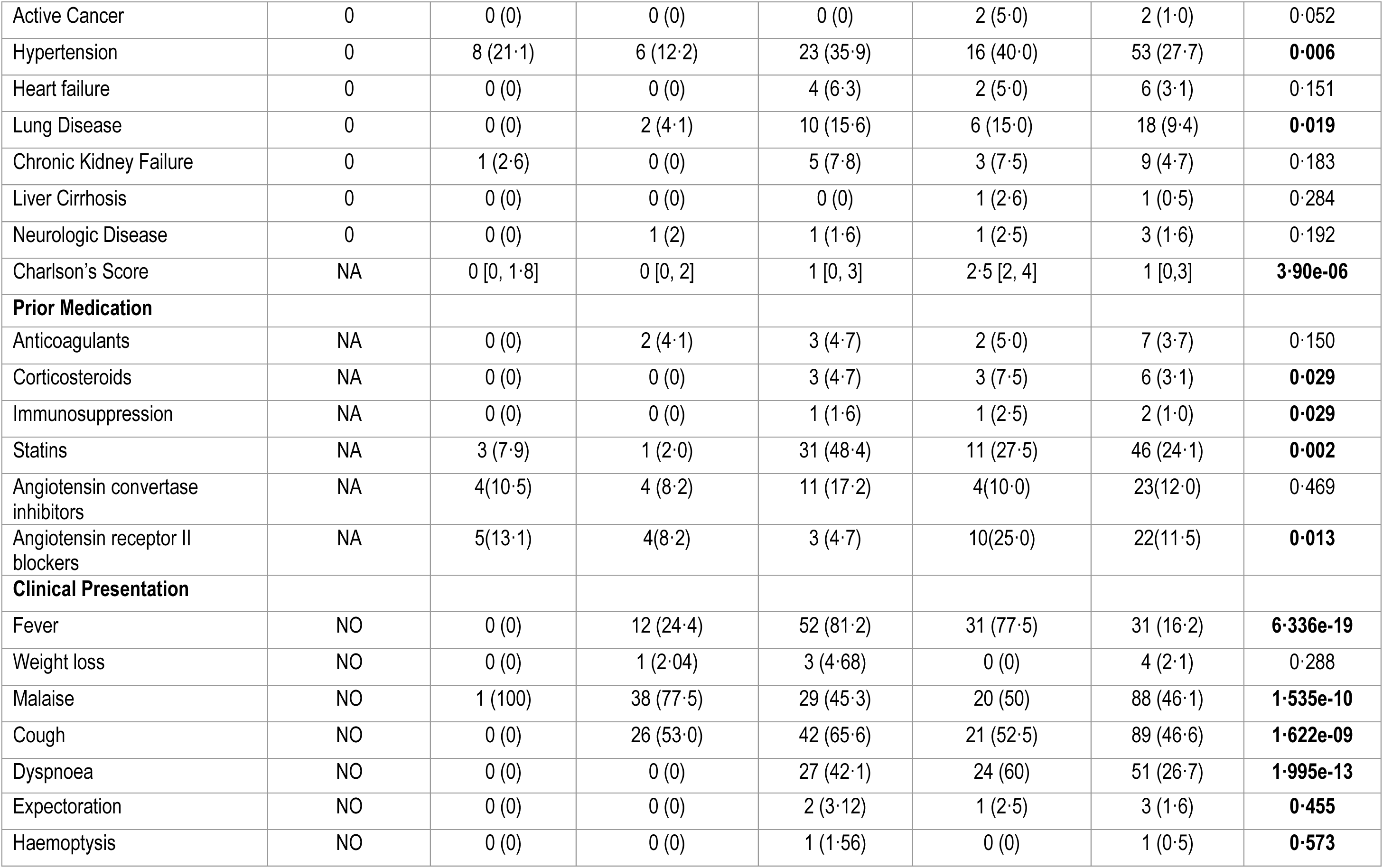
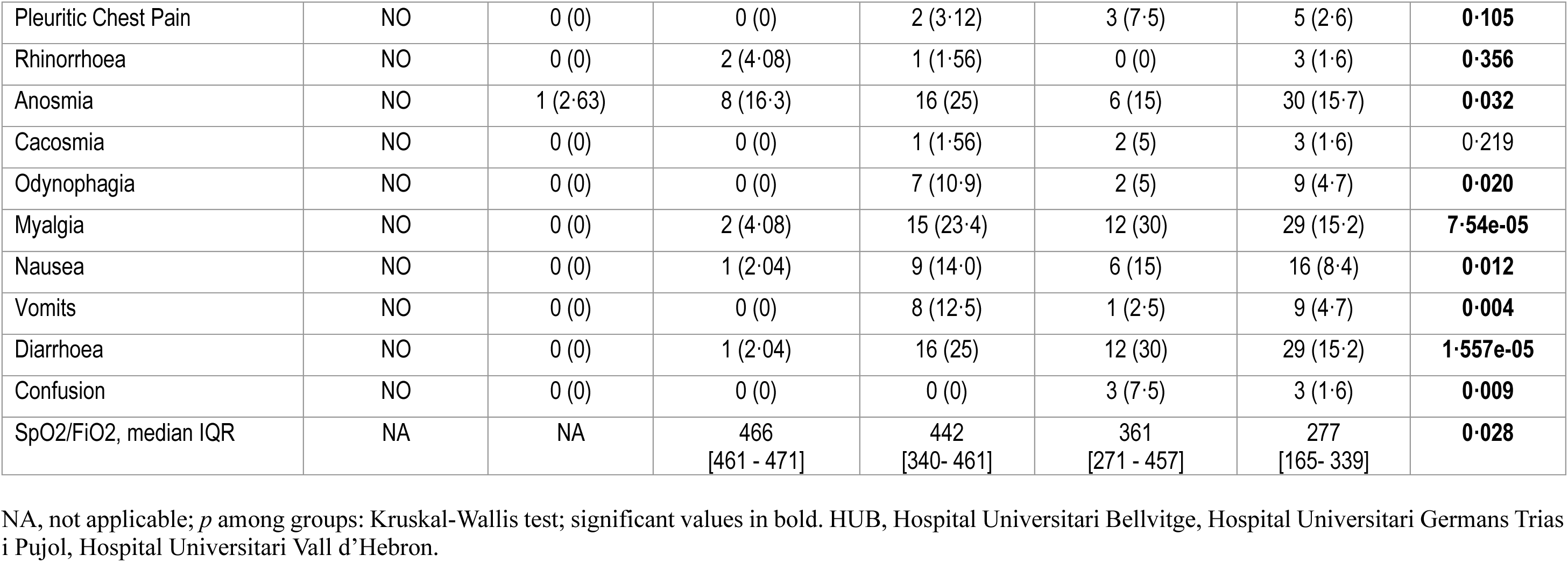
Summary of demographic and clinical data.

**Table 4.** Summary of laboratory data.

|  | Control | Asymptomatic | Mild | Moderate | Severe |  |
| --- | --- | --- | --- | --- | --- | --- |
| Patients | 44 | 38 | 49 | 64 | 40 |  |
| Clinical Chemistry | median [IQR] | median [IQR] | median [IQR] | median [IQR] | median [IQR] | p. adjusted |
| Hb | 15·00 [13·50, 16·02] | 14·30 [13·62, 15·07] | 14·20 [13·35, 14·95] | 13·35 [12·35, 14·70] | 13·20 [12·10, 14·22] | 4·33E-05 |
| WBC | 6·30 [5·16, 7·77] | 5·65 [4·55, 6·47] | 4·50 [3·70, 5·90] | 5·28 [4·10, 6·62] | 7·45 [6·07, 12·66] | 1·90E-07 |
| Basophils 10e9/L | 0·00 [0·00, 0·10] | 0·00 [0·00, 0·00] | 0·00 [0·00, 0·00] | 0·00 [0·00, 0·01] | 0·00 [0·00, 0·02] | 6·14E-03 |
| Basophils % | 0·70 [0·50, 0·92] | 0·50 [0·30, 0·70] | 0·50 [0·30, 0·60] | 0·30 [0·20, 0·50] | 0·20 [0·10, 0·30] | 1·90E-07 |
| Eosinophils 10e9/L | 0·10 [0·10, 0·20] | 0·10 [0·00, 0·10] | 0·10 [0·00, 0·10] | 0·00 [0·00, 0·02] | 0·00 [0·00, 0·01] | 1·90E-07 |
| Eosinophils % | 2·05 [1·67, 3·20] | 1·45 [0·83, 2·30] | 1·20 [0·55, 1·85] | 0·20 [0·00, 0·70] | 0·00 [0·00, 0·10] | 1·90E-07 |
| Lymphocytes 10e9/L | 1·90 [1·50, 2·42] | 1·95 [1·40, 2·38] | 1·60 [1·30, 1·85] | 1·00 [0·86, 1·42] | 0·86 [0·50, 1·20] | 1·90E-07 |
| Lymphocytes % | 30·40 [25·65, 33·95] | 35·70 [30·65, 41·08] | 34·70 [28·00, 41·90] | 21·30 [13·10, 26·20] | 9·40 [5·25, 16·95] | 1·90E-07 |
| Monocytes 10e9/L | 0·50 [0·40, 0·60] | 0·50 [0·32, 0·70] | 0·50 [0·40, 0·60] | 0·45 [0·30, 0·60] | 0·40 [0·30, 0·62] | 2·79E-01 |
| Monocytes % | 7·80 [6·97, 9·05] | 8·45 [7·30, 11·42] | 10·20 [8·90, 13·35] | 8·80 [5·55, 11·10] | 5·05 [3·77, 7·82] | 1·90E-07 |
| Neutrophils 10e9/L | 3·40 [2·95, 4·85] | 2·75 [2·20, 3·50] | 2·30 [1·80, 3·00] | 3·54 [2·69, 4·72] | 7·50 [4·56, 11·44] | 1·90E-07 |
| Neutrophils % | 58·50 [53·48, 62·30] | 50·80 [46·10, 57·95] | 49·40 [44·60, 58·50] | 68·85 [58·68, 80·03] | 82·75 [78·05, 89·25] | 1·90E-07 |
| Platelets 10e9/L | 218·50 [188·50, 255·25] | 197·50 [172·50, 234·75] | 197·00 [169·50, 214·50] | 214·00 [164·50, 278·25] | 236·00 [181·25, 327·00] | 2·42E-02 |
| C Reactive protein mg/dL (0·03–0·5 mg/dL) | ND | 0·23 [0·15, 0·95] | 0·39 [0·20, 1·46] | 6·18 [2·41, 10·76] | 11·91 [7·55, 19·55] | 1·90E-07 |
| Calprotectin | ND | 1·50 [0·90, 1·96] | 1·17 [0·91, 1·78] | 4·80 [3·01, 9·00] | 9·78 [6·14, 22·76] | 1·90E-07 |
| D-dimer (0–243 ng/mL) | ND | 306·00 [251·00, 408·00] | 326·00 [213·00, 517·00] | 309·00 [250·00, 612·00] | 417·00 [266·50, 958·25] | 1·53E-01 |
| Ferritin (25–400 ng/mL) | ND | 120·50 [36·75, 283·50] | 173·00 [66·50, 316·75] | 415·00 [215·50, 1130·00] | 738·00 [508·50, 1383·75] | 1·90E-07 |
| LDH (120–246 IU/L) | ND | 171·00 [157·25, 199·25] | 174·00 [158·50, 201·25] | 250·00 [197·50, 331·50] | 389·00 [304·50, 480·00] | 1·90E-07 |
| ALT 10–49 IU/L) | ND | 20·00 [14·00, 32·00] | 21·00 [15·00, 33·00] | 36·00 [17·00, 61·50] | 35·00 [21·75, 49·25] | 1·13E-03 |
| AST (8–34 IU/L) | ND | 23·00 [18·25, 27·75] | 25·00 [21·00, 32·25] | 39·00 [24·50, 55·00] | 39·00 [31·50, 55·00] | 1·90E-07 |
| Total Bilirubin (0·3–1·2 mg/dL) | ND | 0·42 [0·31, 0·53] | 0·46 [0·36, 0·62] | 0·50 [0·41, 0·70] | 0·53 [0·36, 0·58] | 1·99E-02 |
| Triglycerides (43 – 200 mg/dL) | ND | 85·00 [69·50, 121·25] | 107·00 [74·00, 133·00] | 131·00 [100·00, 158·50] | 134·50 [89·75, 201·75] | 3·88E-04 |
| Creatinine (0,67 - 1·17 mg/dL) | ND | 0·82 [0·71, 1·02] | 0·82 [0·69, 0·99] | 0·81 [0·67, 0·94] | 0·82 [0·71, 0·94] | 9·60E-01 |
| Urea (17–42 mg/dL) | ND | 29·40 [25·35, 35·70] | 30·00 [25·80, 36·60] | 33·00 [26·38, 48·45] | 48·90 [39·90, 57·50] | 1·90E-07 |
| Fibrinogen ((2·39–6·1 g/L) | ND | 4·63 [3·96, 5·26] | 4·47 [3·91, 4·95] | 6·00 [4·00, 7·00] | 6·00 [5·00, 7·00] | 3·54E-06 |
| Prothrombin Time INR | ND | 1·10 [1·04, 1·13] | 1·10 [1·05, 1·16] | 1·02 [0·98, 1·12] | 1·07 [1·00, 1·13] | 4·82E-02 |
| Anti-SARS-CoV-2 Serology |  |  |  |  |  |  |
| Anti-SARS-CoV-2 Mpro IgM | 33·26 [22·03, 47·63] | 39·10 [24·23, 58·06] | 33·31 [25·12, 68·87] | 76·69 [28·52, 202·40] | 71·48 [27·81, 246·80] | 1·35E-03 |
| Anti-SARS-CoV-2 NP IgM | 19·79 [10·30, 36·46] | 22·87 [12·06, 40·19] | 20·17 [11·95, 52·20] | 49·58 [17·15, 153·28] | 44·92 [19·82, 191·47] | 5·66E-04 |
| Anti-SARS-CoV-2 Spike IgM | 11·26 [7·73, 16·63] | 18·14 [11·35, 23·69] | 18·27 [10·64, 50·39] | 195·79 [32·00, 419·62] | 166·40 [19·37, 597·09] | 1·90E-07 |
| Anti-SARS-CoV-2 RBD IgM | 28·30 [18·08, 50·42] | 31·68 [18·74, 51·56] | 34·24 [22·47, 57·83] | 27·30 [18·18, 56·10] | 28·44 [13·35, 67·82] | 6·45E-01 |
| Anti-SARS-CoV-2 Mpro IgG | 104·62 [79·80, 140·57] | 131·61 [98·43, 213·75] | 141·66 [103·37, 251·88] | 2366·32 [218·92, 8961·47] | 3802·74 [255·20, 17061·04] | 1·90E-07 |
| Anti-SARS-CoV-2 NP IgG | 87·82 [52·94, 147·42] | 108·90 [67·80, 299·94] | 119·68 [65·33, 219·93] | 1523·34 [213·28, 6132·72] | 1675·92 [198·03, 14891·76] | 1·90E-07 |
| Anti-SARS-CoV-2 Spike IgG | 352·43 [172·59, 519·99] | 482·34 [413·13, 781·45] | 363·83 [287·60, 674·92] | 1182·96 [479·80, 2882·23] | 1409·81 [675·78, 3069·88] | 1·90E-07 |
| Anti-SARS-CoV-2 RBD IgG | 151·80 [99·39, 189·06] | 166·19 [141·64, 197·79] | 169·66 [127·47, 230·83] | 174·49 [124·02, 258·31] | 206·29 [150·35, 267·33] | 2·09E-02 |
| Anti-SARS-CoV-2 Mpro IgA | 63·42 [53·66, 78·82] | 73·47 [52·11, 156·64] | 82·32 [53·31, 147·56] | 1863·08 [141·85, 7094·48] | 1950·15 [170·74, 21127·41] | 1·90E-07 |
| Anti-SARS-CoV-2 NP IgA | 47·75 [38·01, 66·72] | 60·97 [37·51, 128·13] | 65·71 [39·42, 145·72] | 2233·86 [139·40, 8282·62] | 1494·22 [165·56, 22643·54] | 1·90E-07 |
| Anti-SARS-CoV-2 Spike IgA | 89·72 [71·30, 133·01] | 99·82 [64·31, 290·57] | 219·47 [88·11, 307·64] | 2401·77 [267·93, 10810·81] | 3780·36 [478·96, 9936·22] | 1·90E-07 |
| Anti-SARS-CoV-2 RBD IgA | 97·75 [89·98, 110·80] | 101·33 [83·12, 117·93] | 101·37 [91·10, 122·64] | 108·62 [89·57, 152·74] | 101·66 [89·19, 148·74] | 2·96E-01 |
| Cytokines and chemokines |  |  |  |  |  |  |
| IFN-alpha | 0·00 [0·00, 0·20] | 3·52 [0·30, 25·55] | 15·75 [2·17, 37·62] | 15·25 [2·63, 33·75] | 2·05 [0·05, 15·85] | 8·68E-07 |
| IFN-gamma | 0·79 [0·64, 0·97] | 2·28 [1·08, 6·73] | 3·04 [1·69, 5·01] | 9·61 [3·04, 24·89] | 3·50 [2·10, 9·84] | 1·90E-07 |
| TNF-alpha | 12·41 [9·91, 15·28] | 20·95 [16·53, 24·30] | 21·94 [18·42, 27·83] | 21·30 [16·84, 30·80] | 21·57 [18·75, 27·06] | 1·90E-07 |
| IL-6 pg/ml | 3·06 [2·33, 4·46] | 3·44 [2·67, 9·15] | 5·12 [3·46, 15·40] | 33·85 [19·82, 56·38] | 37·20 [16·10, 107·02] | 1·90E-07 |
| IL-1RA pg/ml | 365·02 [269·41, 504·56] | 581·58 [443·75, 922·75] | 940·33 [539·00, 1548·07] | 1727·37 [1281·84, 2839·75] | 2190·32 [1377·82, 3004·16] | 1·90E-07 |
| IL-7 pg/ml | 13·89 [10·13, 16·30] | 14·15 [10·70, 18·85] | 15·57 [10·61, 18·24] | 21·90 [13·12, 32·41] | 39·30 [25·55, 55·57] | 3·58E-07 |
| IL-12p70 pg/ml | 1·16 [0·29, 2·75] | 2·45 [1·68, 2·84] | 2·13 [1·18, 2·93] | 1·77 [1·24, 2·11] | 1·60 [1·05, 2·15] | 2·03E-02 |
| IL-10 pg/ml | 2·98 [1·85, 3·43] | 4·46 [3·80, 8·32] | 5·48 [3·63, 8·25] | 12·10 [8·74, 17·90] | 19·20 [11·75, 23·15] | 1·90E-07 |
| IL-13 pg/ml | 6·74 [4·06, 11·20] | 8·60 [3·32, 16·40] | 9·33 [6·43, 15·65] | 7·20 [5·62, 18·62] | 8·75 [5·18, 15·72] | 2·95E-01 |
| IL-15 pg/ml | 2·83 [2·09, 4·00] | 3·53 [2·87, 4·98] | 4·98 [3·50, 6·39] | 5·35 [4·19, 7·35] | 5·52 [4·61, 6·79] | 3·58E-07 |
| IL-17A pg/ml | 1·17 [0·69, 2·00] | 2·41 [1·71, 3·14] | 2·76 [2·03, 3·90] | 2·57 [2·10, 4·54] | 2·58 [1·57, 3·91] | 6·67E-05 |
| IL-2 pg/ml | 0·15 [0·08, 0·30] | 0·30 [0·22, 0·45] | 0·44 [0·32, 0·64] | 0·84 [0·51, 2·04] | 0·76 [0·48, 1·03] | 1·90E-07 |
| IL-4 pg/ml | 0·66 [0·53, 0·78] | 0·50 [0·35, 0·61] | 0·58 [0·45, 0·79] | 0·72 [0·47, 1·04] | 0·70 [0·57, 0·86] | 2·42E-02 |
| GM-CSF pg/ml | 1·62 [1·26, 2·04] | 3·10 [2·29, 3·71] | 2·32 [1·45, 3·04] | 2·13 [1·58, 3·37] | 2·98 [1·71, 4·07] | 4·38E-04 |
| TGF beta-1 pg/ml | 114·50 [73·12, 151·25] | 163·00 [104·00, 196·75] | 169·50 [133·00, 229·25] | 125·00 [47·15, 172·50] | 188·00 [103·50, 289·50] | 3·52E-03 |
| CCL2 pg/ml | 438·01 [338·12, 556·64] | 542·00 [423·25, 675·00] | 613·00 [488·55, 919·00] | 653·00 [486·65, 866·25] | 883·00 [489·00, 1576·64] | 1·19E-03 |
| CXCL10 pg/ml | 133·50 [119·94, 165·00] | 466·00 [290·50, 930·00] | 878·00 [584·75, 1478·25] | 1742·91 [1177·00, 2278·00] | 2371·00 [1645·33, 3604·50] | 1·90E-07 |
| GRANB pg/ml | 15·60 [11·30, 18·80] | 33·30 [26·35, 46·75] | 42·20 [26·92, 64·93] | 43·35 [26·00, 65·07] | 36·60 [27·55, 51·55] | 1·90E-07 |

### ANALYSIS OF THE ANTIBODY RESPONSE TO THE SARS-CoV-2 PROTEINS

#### The time course profiles correspond to a recall immune response and are associated with severity but not with the presence of Type-1 IFN autoantibodies

We analysed the serological response to Mpro, NP, Spike (S), and Spike RBD (RBD) in 473 samples from 190 patients and 44 controls. The values were standardised using the control group’s IQR3.

In period DFSO1, the antibody response showed significant differences in control versus all severity group patients, except for anti-RBD (Fig. 2a and b). Interestingly, the only significant IgM response during this period was to the Spike protein. The differences became more evident in the following DFSO periods when the response to RBD and IgM isotype antibodies became detectable (see Supplemental Figure S1a–d). Although the highest responses were initially observed in moderate patients, they resembled those of severe patients during the follow-up. Responses were also detected in asymptomatic patients but were only significant for anti-S IgG.

**Figure 2.**
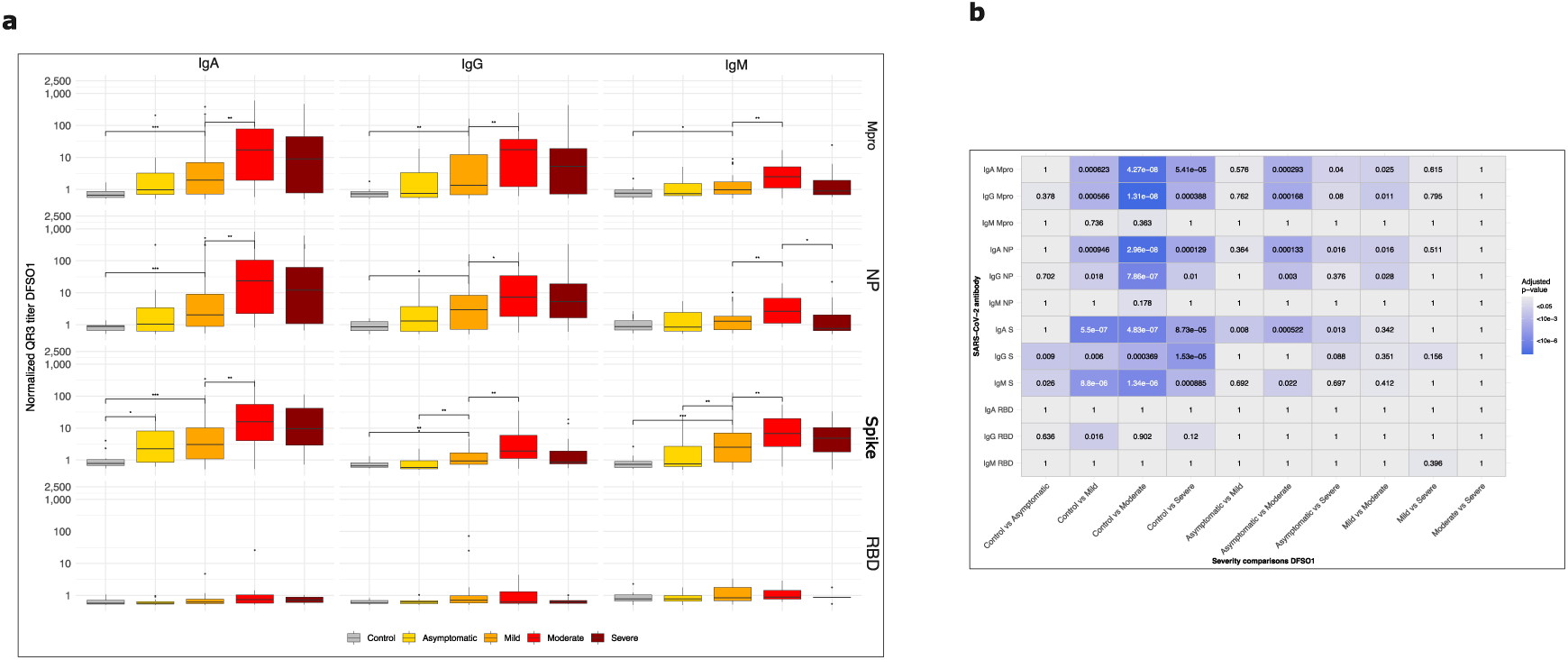
Pairwise comparison of antibody response to SARS-CoV-2 proteins in the different groups of patients. (**a**) The plots correspond to the 0-7 days from symptom onset (DFSO1); only significant differences among adjacent boxplots and controls are annotated. Median ± IQR. (b) *p* values for each possible comparison from a); cells with p values >0.05 are not coloured. Paired Wilcox test, adjusted *p* values. The corresponding graphics for periods DFSO2 and DFSO3 are in Supplemental Figure 1.

The antibody response was more prolonged in patients with more severe symptoms with a considerably later peak response, especially for anti-RBD and IgG isotype antibodies (Fig. 3a and b). To better visualise antibody response trajectories, we plotted severity-stratified data using LOESS smoothed regression that generated curves summarising each isotype’s antibody response (Fig. 3c). Two key observations emerged: 1) IgM responses did not appear before IgA or IgG, neither initially nor at the peak of the response. 2) Antibody levels decreased in mild patients between 13 and 16 days but only after 30 and 47 days for moderate and severe patients.

**Figure 3.**
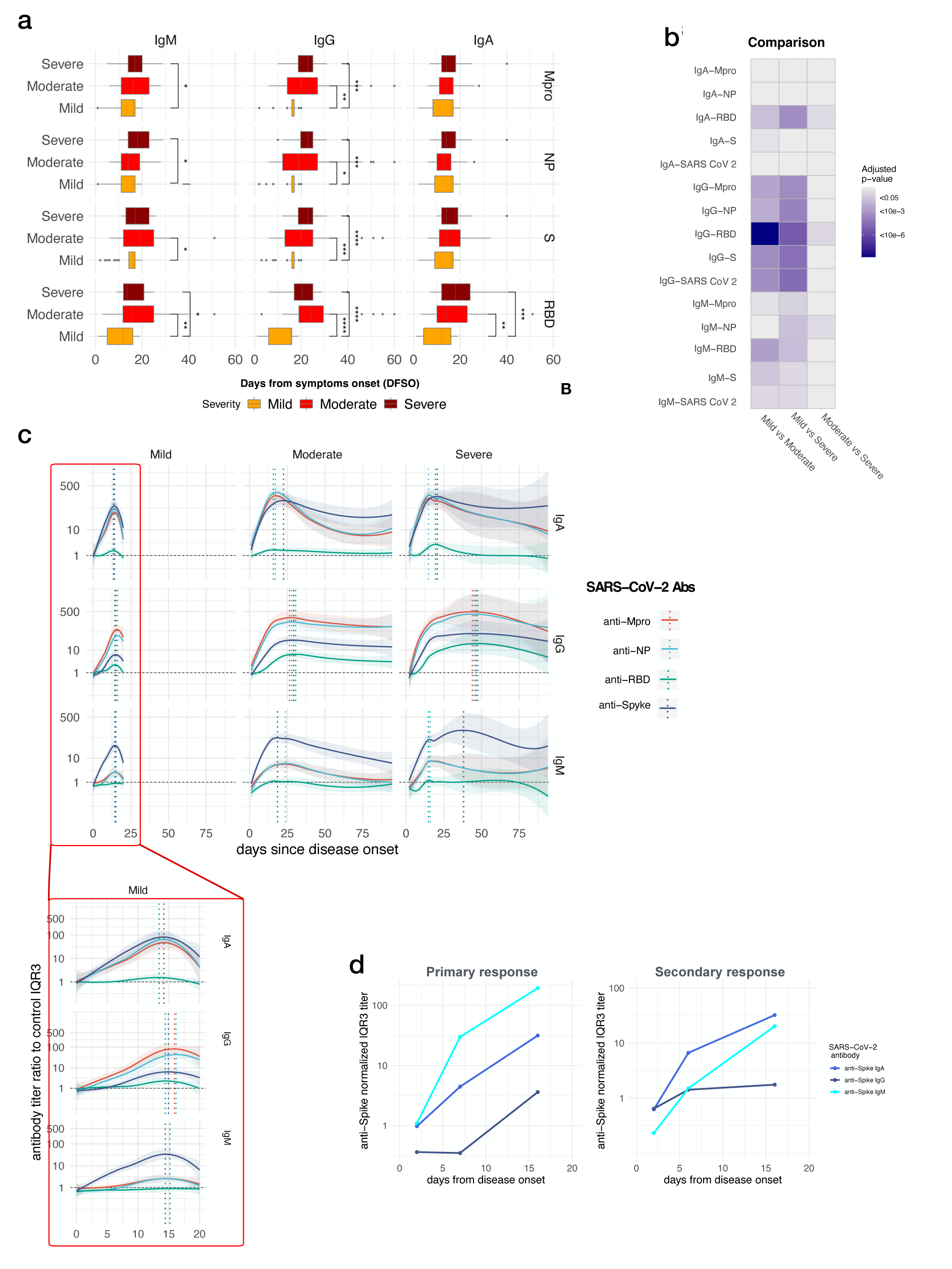
Trajectories of SARS-CoV-2 antibody levels. (a) The peak levels of the 88 cases from which we collected three samples during the initial 60 days are plotted against time. There’s a noticeable delayed peak for moderate and severe cases compared to mild cases. Median ± IQR. (b) A heatmap summarises the significance level of the comparisons, highlighting clustering in the IgG category and the comparison between mild vs. severe and mild vs. moderate cases. (c) LOESS regression curves with 95% confidence intervals of the normalised antibody titres of 392 samples from 190 COVID-19 cases are shown. The vertical dotted lines represent the maximal titre, and the horizontal lines represent the established normal level for data normalisation. In the red box inset, the time scale has been zoomed to visualise the trajectories during the initial 20 days. It is evident that the first response is IgA, followed by IgG and IgM. The responses have already started to decrease at DFSO 13-15 days. The rise of the IgM antibody regression curve never precedes the other isotype curves in moderate or severe patients. The maximal IgG titre is reached between 43 and 47 days for moderate and severe patients. It’s also noteworthy that the predominant antigen for IgM isotype antibodies is Spike, while IgA, NP, Mpro, and NP predominate over Spike for IgG. Responses to RBD were predominantly IgG. (d) Representative patients for primary and secondary responses; only responses to Spike have been represented.

This lack of an IgM response to SARS-CoV-2 preceding the IgG and IgA responses, as expected in an immune response to a new pathogen, suggested that in most patients, the response was, at least in part, a recall response. However, as in a LOESS regression, a few cases with a primary response can be missed, we looked for primary responders by selecting patients who, during the initial seven days, had a normalised level of antibody above 1·5 for the IgM isotype and below 1·5 for the IgG or IgA isotype.

The sum of Mpro, NP, and S antibodies for each isotype and patient was used for this filter. Eight out of 139 cases (5·8%) were identified. They corresponded to four mild and four asymptomatic patients, all from the primary care sub-cohort, none requiring hospital admission (age 52 range 29–71, five females, and three males). Representative profiles are shown in Fig. 3d. These primary responders developed good IgG and IgA antibody responses at period DFSO2 (data not shown). The interpretation of these results is that in this COVID-19 cohort, the majority develop a recall type of immune response to SARS-CoV-2 proteins.

An expected feature of a response to a new pathogen is a coordinated immune response to the pathogen’s different antigens. In our cohort, the responses to the SARS-CoV-2 Mpro and NP proteins are coordinated, but not the response to S and especially to the RBD (Supplemental Figure S2. blue boxes). As the receptor binding domain (RBD) of SARS-CoV-2 differs significantly from other circulating coronaviruses, a delayed and slow response to RBD would indicate the need for a primary immune response distinct from the response to other SARS-CoV-2 proteins.

A reported observation is that in COVID-19, the antibody response titre correlates positively with the disease’s severity. In our cohort, maximal antibody titres were indeed associated with severity on the five-point scale (control, asymptomatic, mild, moderate, and severe, Fig. 4a and b). This correlation was also found when stratified by WHO scores (Supplemental Figure S3a and b).

**Figure 4.**
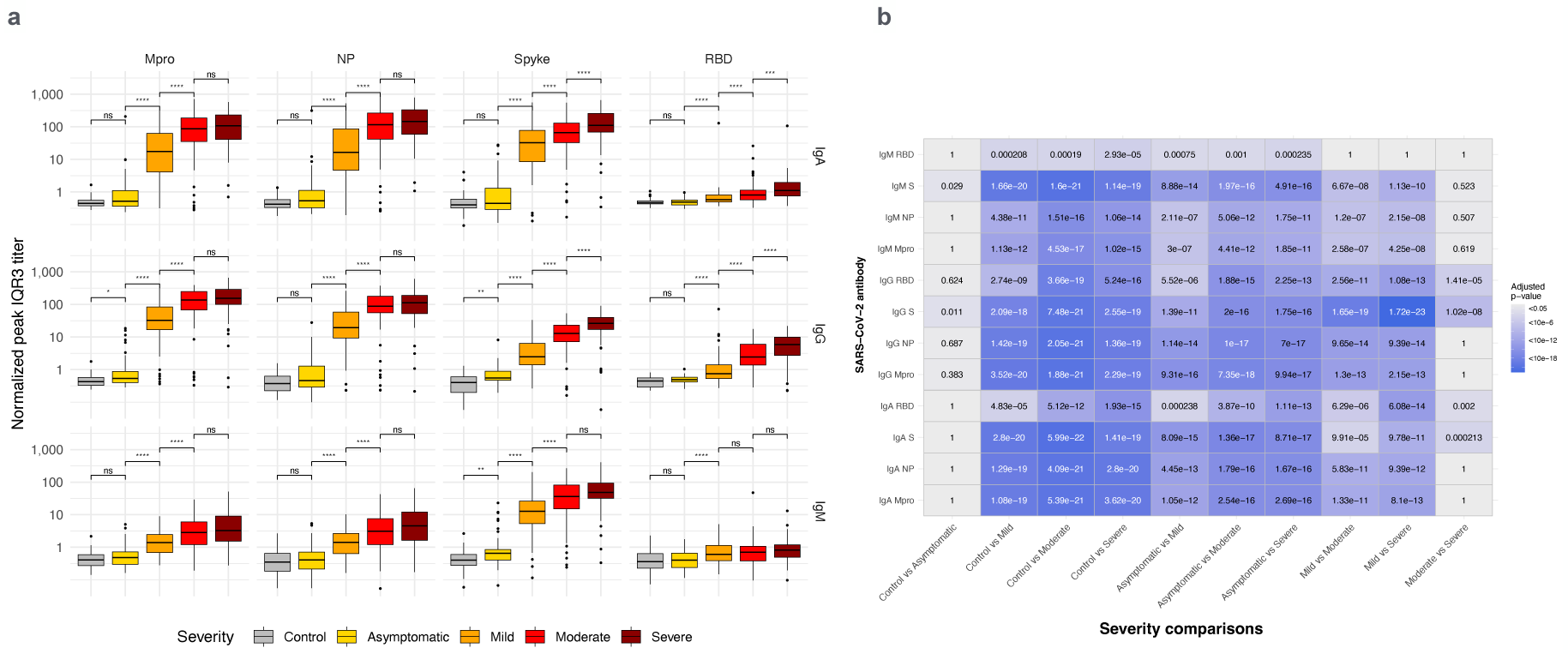
The intensity of the antibody response increases with severity. (A) Maximal antibody levels were compared for the clinical cases with three serological measurements plus control and asymptomatic cases. Notice how the antibody levels significantly increase with the severity. Median ± IQR. (B) Heatmap summarising plot comparisons, Pairwise Wilcoxson test. See supplemental Figure S3 for the same analysis but with the WHO eight-point scale.

The interpretation of the association of severity with the serological response is that antibodies do not protect against severity during the early stages of COVID-19, notwithstanding their protective role as RBD-neutralizing antibodies against re-infection. ^29^

As anti-type I IFN autoantibodies have been associated with the severity of COVID-19 ^6^, we measured anti-IFN-alpha and anti-IFN-omega autoantibodies. Only 10 of the 232 (4·9%) COVID-19 cases and two of the 44 controls (4·5%) were positive. Of these ten positive cases, one was asymptomatic, three moderate, three severe, and one deceased; three were female, and four were males (including the deceased). This small number of positive cases indicated that anti-type I IFN autoantibodies were not an essential determinant of severity in this cohort and did not interfere significantly with the analysis and its interpretation.

#### Cytokine profiles are consistent with a recall immune response to SARS-CoV-2

To identify which cytokines were driving the humoral response, we analysed the correlations of cytokines with the antibody titres at each period and severity group. The most interesting observation is the significant negative correlation of IFN-γ, especially with the serological response in moderate patients at DFSO2; it is known that there is a mutual inhibitory effect of IFN-γ/IL-2 and IL-4/IL-13 in the initial polarisation of the immune response, which may explain these results ^30^ (Supplemental Figure S4). The positive correlation of IL-7 with antibody titres in severe and moderate patients is probably due to IL-7 being secreted to compensate for lymphopenia, which correlates negatively with antibodies to SARS-CoV-2. ^31^ Overall, the pattern of cytokines with peaks at DFSO2 for mild and at DFSO3 for moderate and severe cases suggest a mixture of early and late secondary immune responses.

### ANALYSIS OF THE BLOOD CELL POPULATIONS IN COVID-19 PATIENTS

#### Changes in the main leukocyte populations

As reported, neutrophils were relatively expanded in COVID-19 patients, and lymphocytes were reduced. ^8^ To better analyse B and T cell populations, we have used the absolute number and the proportion of a subset within each lymphocyte’s main compartment. Low T lymphocytes, monocytes, and NK cells were detected during the initial DFSO1 period. The most striking change was in plasma/plasmablast cells, which increased in all severity categories (Supplemental Figure S5). The spectral flow cytometric analysis of total blood resolved 46 populations further split into 197 clusters. For the interpretation, three degrees of resolution were considered: low (7 subsets), medium (46 subsets), and high (197 clusters). Only those contributing to discern between a primary and a recall response are included in this report.

#### Trajectories of plasmablast subsets show features of a recall response

Plasmablasts and plasma cells, from here on PBs, as defined by CD38 +, CD27+, HLA-DR++, sIg, and variable CD19, were split into four main clusters: IgA+, IgG+, IgM+, and sIg^-^ and three low-abundance clusters: IgM+IgA+, IgM+IgG+, and long-life plasma cells (LLPC), as seen in the uniform manifold approximation and projection plot (UMAP) (Fig. 5a and b).

**Figure 5.**
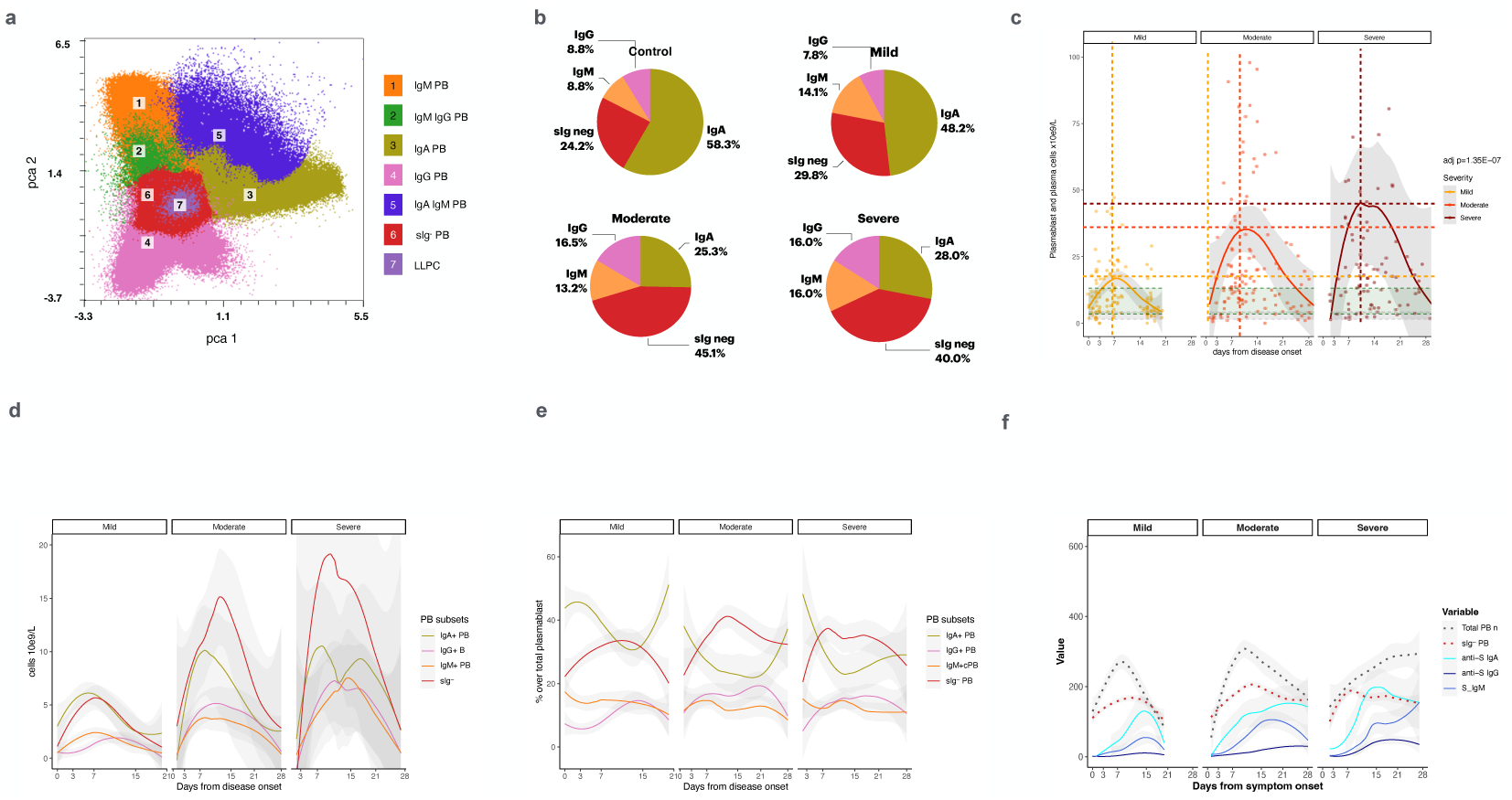
Plasmablast (PB) expansion within the WBC, lymphocyte populations, and subset analysis. (**a**) UMAP of plasmablast clusters; (b) Distribution of plasmablast among the four main subpopulations, IgA+, IgG+, IgM+, and sIg^-^ in controls and the three severity categories. The expansion of PB is due to the rise in the number of sIg^-^ PBs. (c) LOESS trajectories of total plasmablasts during the initial 28 days by severity categories. The horizontal dashed lines indicate the different levels of the maximal number of cells. The vertical dashed lines indicate the day the maximal level is reached for each category, highlighting the remarkable differences in their respective trajectories. The magnitude of PB’s absolute expansion can be appreciated (d and e). The comparison of PB subset trajectories shows that their expansion is due to the sIg^-^ PB subset. (f) Composite trajectory plot showing total PBs and sIg-PB trajectories and normalised antibody titres to SARS-CoV-2 S proteins. Values in the y-axis are cell numbers or normalised antibody titres; cell numbers were transformed to place them in the same range as serological titres.

The number of PBs rose rapidly in moderate and severe patients, surpassing at DFSO1 day three the maximal levels of mild patients at day seven; their peak exceeded the peak in mild patients by a factor of two and three, respectively (Fig. 5c). In some severe patients, PBs made up to 60% of lymphocytes (Supplemental Figure S6a). From the trajectories, it is inferred that IgA+ PB was the dominant subset during the incubation period and was replaced by sIg-PB on DFSO day 3 (Fig. 5d and e). The comparison of total PB cells and antibody titre trajectories suggests that PBs cells are the source of at least some of these antibodies to SARS-CoV-2 (Fig. 5f). The correlation between sIg^-^ PB number and anti-SARS-CoV-2 antibodies in mild patients at DFSO1 supports this trend (Supplemental Figure 6b). A negative correlation of PBs with Th1 cytokines IFN-gamma and IL-2 and a positive correlation with IL-10 and IL-7 is consistent with an ongoing humoral immune response (Supplemental Figure S7).

Interpretation: The first IgA plasmablasts probably originate from upper airway mucosa secondary lymphoid organs (SLO). Given the magnitude, dynamics, and short incubation time of COVID-19 ^32^, they are more likely to result from memory B cells specific to the cross-reactive CCCV virus than from a fast primary response.

#### Trajectories of the B lymphocyte subset in COVID-19 patients are consistent with a recall response

Flow cytometry identified 32 clusters of B lymphocytes summarised in 13 subsets and eight unclassified minor clusters (Fig. 6 a and Supplemental Figure S8). The total B cell trajectory differs from that of leukocytes and T cells; still, with time, their shifts show parallelism (Fig. 6b). Subset trajectories are very variable (Fig. 6d–h). Switched and memory subset trajectories differed markedly among severity groups and also when compared by period and severity (Supplemental Table 2).

**Figure 6.**
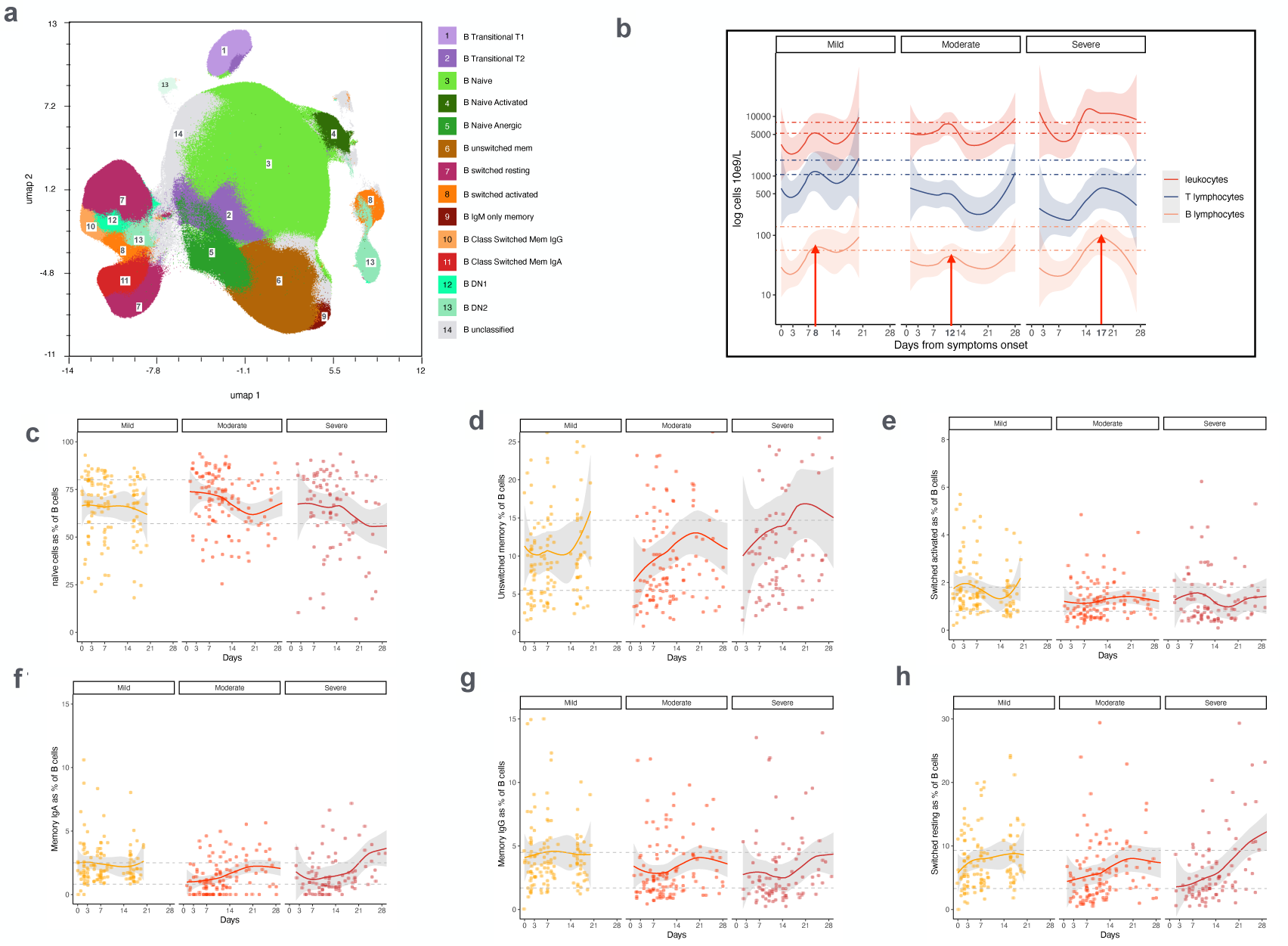
Trajectories of B cell subsets. (a) UMAP of B cells displays the distribution of the 13 subpopulations summarising 32 clusters. (b) LOESS trajectory of leucocytes, T and B lymphocytes counts for the three categories of clinical COVID-19. In plots c-h, the samples are mild 130, moderate 134, and severe 89. The y-axis represents the percentage of total B cells. (c) Naïve; (d) unswitched memory; (e) Switched activated; (f) Memory IgA; (g) Memory IgG; (h) switched resting B cells. The dashed lines indicate quartiles 1 and 3 of the distribution of the values in the control population for each subpopulation. By comparing each population (c-h) with (b), it is noticeable that the circulating B cell population is less reduced compared to total lymphocytes and that recovery is earlier in the less severely ill patients.

The B cell subset correlation analysis with the serological response revealed a distinct pattern. In mild patients, the correlations are positive during the initial eight days (DFSO1), significant for B cell subsets involved in the early phase of the response, i.e., transitional, naïve activated, IgM-only memory, and CD24++ immature IgG memory (Fig. 7a and Supplemental Figure S9 for high-resolution B cell clusters). This correlation is clearly detected in moderate and severe at DFSO2. Consistently, the trajectories of memory and switched subsets show a delay in the regression curves (Fig. 7b) and when analysed by periods in the ring doughnuts plot (Fig. 7c).

**Figure 7.**
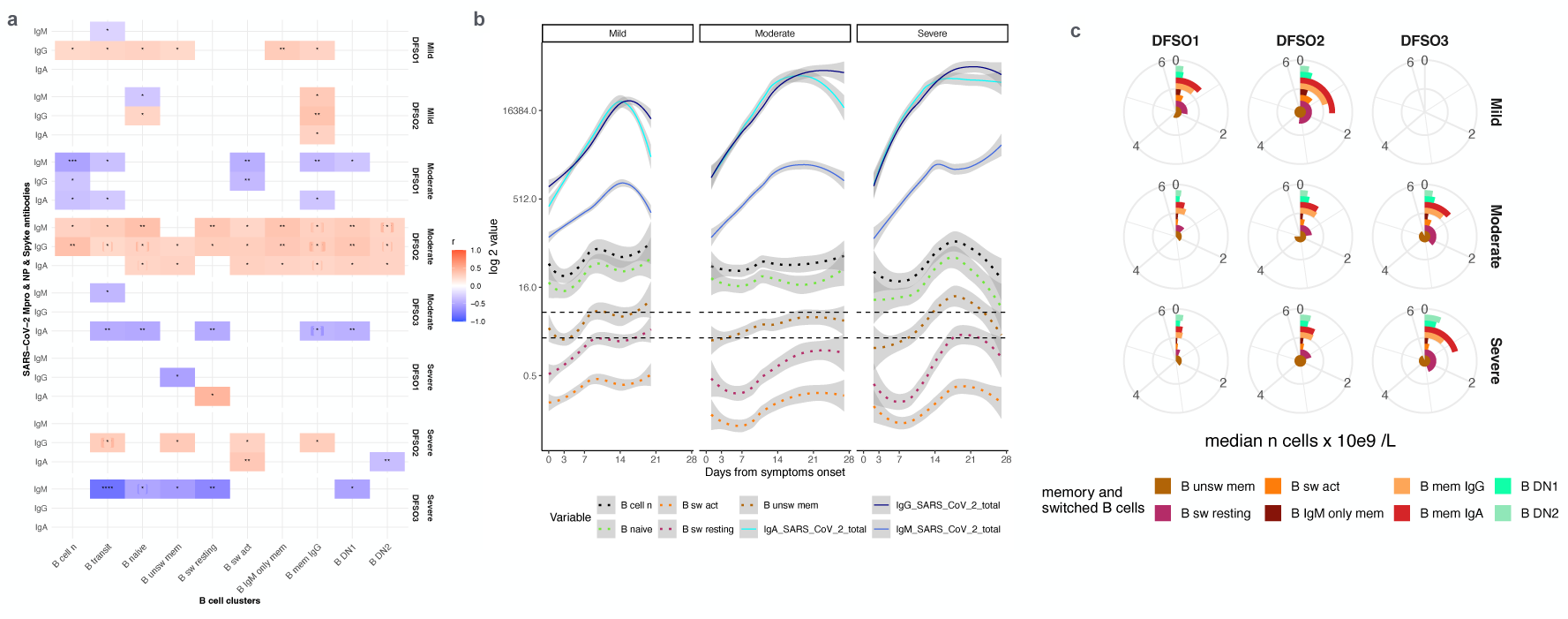
Trajectories of B cell subsets associated with a recall immune response. (**a**) Heatmap summarising the correlation (Spearman) of B cell subsets and serological response by severity and follow period. DFSO, days from symptoms onset. DFSO1 0-7. DFSO2 8-20, DFSO3 21-107. (b) LOESS regression trajectories comparing B cell subsets and antibodies to SARS-CoV-2 proteins. (c) Ring doughnut charts comparing the distribution of the B cell subset along the follow-up period by severity. See text for details. Notice that the inner circles represent the most abundant subpopulations.

Interpretation: The overall cell dynamic is consistent with a recall rather than a primary response. The capture of cross-reactive antigens by CCCV memory B cells would delay the recruitment of naïve B cells in moderate and severe cases. A fresh, specific response to SARS-CoV-2 would be initiated only when a large amount of antigen reaches the SLO.

#### Trajectories of cTQ cells in COVID-19 patients are consistent with a recall response

Circulating Tfh (cTfh) reflects the activity of Tfh cells in the SLO. ^33–35^ It correlates with antibody response, but the trajectory differs from total T cells or typical effector memory CD8 cells (Fig. 8a). cTfh, as Tfh, can be classified as cTfh-naïve, cTfh1, cTfh17, cTfh2, and cTfh1 activated, ^35^ as identified in the T cells UMAP (Fig. 8b). All clusters were significantly higher in asymptomatic and mild than in moderate and severe (Fig. 8c). In severe patients, all T cell subsets contract. Still, Tfh is the significantly more contracted subset. This association of cTfh contraction with severity was not explained by age or sex in the multivariate model. Notably, despite the low number of cTfh cells, the rise in antibodies to SARS-CoV-2 is earlier and faster in moderate and severe patients than in mild patients, suggesting that they are produced by memory B cells that require less Tfh help. CXCR5, the defining cTfh marker, was higher in asymptomatic and mild patients than in moderate and severe patients, indicating recent egress from SLO in these patients (Supplemental Figure S10a). The high positive correlations in the cTfh clusters to serological response analysis confirm that Tfh participates in the antibody response (Supplemental Figure S10b).

**Figure 8.**
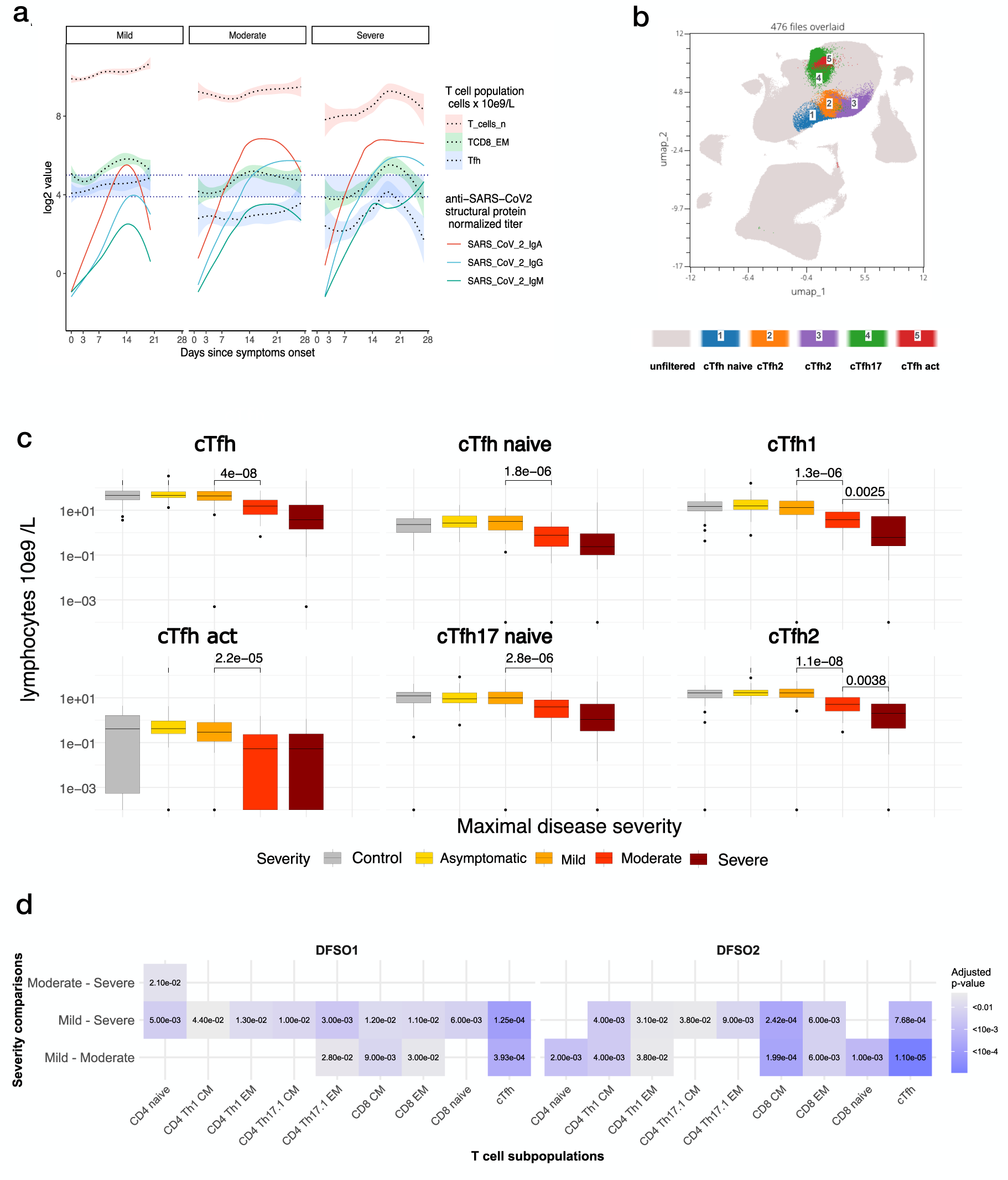
cTfh and clusters. (a) Comparison of LOESS trajectories of total T lymphocyte number, CD8 EM, and cTfh with the LOESS trajectories of normalised antibody to SARS structural proteins. The 95% CI of trajectories is represented; the blue ribbon helps to visualise how cTfh in mild remains consistently above the levels in moderate and severe patient groups. Notice that in moderate and severe, the antibody titres rise despite a lower number of cTfh. Median ± IQR. (b) UMAP shows the distinct cTfh clusters within the T cell distribution. (c Both total Tfh (top left panel) and each cTfh subcluster have significantly different values distributions for mild vs moderate (Wilcoxon test p. adjusted Benjamin-Hochberg). (d) Heatmap showing the pairwise comparison of T cell subsets among the severity groups. cTfh distribution is the most significantly different (Wilcoxon test p. adjusted Benjamin-Hochberg).

Interpretation: The overall pattern of cTfh response aligns with a recall response, but the higher number in mild patients points to some priming to the differential SARS-CoV-2 epitopes in these patients.

#### Nanostring transcriptomic signature

Seventy samples from 33 patients, representative in terms of age, sex, and severity, were selected for transcriptomic profiling. The results were stratified for the analysis in 60 gene groups.

The strong BCR signalling signatures in asymptomatic patients are of interest because other techniques detected only slight changes in this group (Fig. 9). In the Immune memory panel, the stronger signal for CD45RA in asymptomatic and mild patients with the differential pattern in the lymphocyte trafficking highlights the different regulation of the immune responses in asymptomatic and mild vs the hospitalised moderate and severe. It is also remarkable that there is an interferon response signature in the asymptomatic and some mild cases. The results are consistent with an immune response, mainly recall in moderate and severe patients.

**Figure 9.**
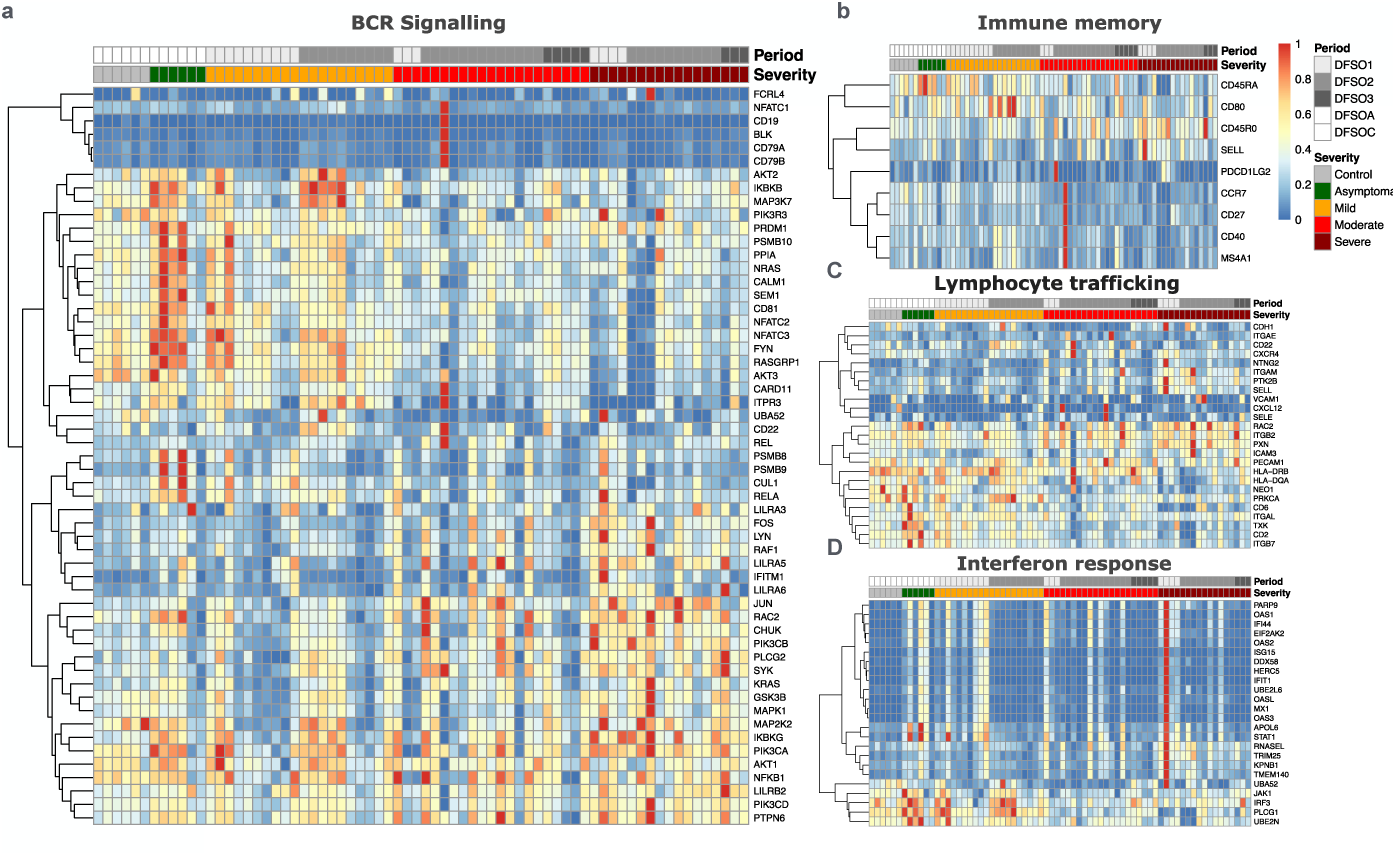
Heatmaps summarising the transcriptomic profile of 33 COVID-19 patients stratified by severity and follow-up periods. (a) BCR signalling gene set; (b) Immune memory gene set; (c) Lymphocyte Trafficking gene set; and (d) Interferon signalling gene set.

The panels of genes associated with myeloid and monocyte cell activation and the expected IL-1 signalling pathway show some of the main differences related to severity (Supplemental Figure S11). These results are similar to many transcriptomic profiles of COVID-19 patients and indicate that our cohort is representative of COVID-19. ^36–38^

## Discussion

At the start of the COVID-19 pandemic, it was appreciated that around 20% of the patients developed pneumonia, many of which required oxygen therapy and intensive care unit admission. Attempts were made to generate predictive algorithms based on immunological variables to help triage patients. These analyses identified several immunotypes;^39^, however, to reach a practical predictive value, algorithms also had to include clinical signs of severity ^8,40^, highlighting the limitations of tools available to identify the immunological determinants of severity. This suggests that not all the relevant immunological variables were taken into account.

One possible immunological determinant of severity is the existence of immunological memory of CCCV antigens that cross-react with SARS-CoV-2 and enhance or interfere with the immune response, the mechanism known as antigenic imprinting (IP) or original antigenic sin. This possibility has been discussed. ^15,41,42^ and there is evidence that most individuals possess antibodies ^16^ and memory T cells ^15,43^ to CCCV that cross-react with SARS-CoV-2 proteins. A recent study established a link between pre-existing immunity to CCCV and COVID-19 severity ^20^. Conceptually, prior immunity to CCCV may be protective or detrimental depending on the localisation of the cross-reactive epitopes, the type of memory, and the coexisting determinants of severity in a given patient. ^18^ Despite this evidence, IP has not become a central paradigm in the pathogenesis of severe COVID-19 because of discrepancies in the effect of pre-existing immunity on the outcome ^22,29^.

There is a lack of studies focused on whether the immune response to SARS-CoV-2 is primary or recall and whether there is an association between the type of response and severity; this is relevant because only a recall response can be subjected to IP / original antigenic sin.

In this paper, we report that the trajectories of antibody responses to SARS-CoV2, when analysed in depth, together with the clinical course and the shifts in plasmablasts, B cell, and cTfh subpopulations indicate that in 131 out of 139 (94·3%) cases, the immune response to SARS-CoV-2 is a recall response. Interestingly the features of recall response were more evident in the two categories of patients that required hospitalisation. The direct implication is that since the immune response to SARS-CoV-2 is a recall response in most cases, IP can be a significant determinant of SARS-CoV-2 infection severity in most COVID-19 cases.

Our study is prospective; clinical data of all cases were curated by their physicians; patients were from the same geographical area and a single health provider (Institut Català de la Salut, (ICS), https://ics.gencat.cat). Spectral flow cytometry was conducted on fresh blood samples rather than on cryopreserved cells; cytokines were measured using Biotechne® ELLA microfluidic assays, a platform selected for use in the HUVH clinical laboratory because of its robustness and in use since 2018; transcriptomic data were obtained using Nanostring®, which does not include a PCR-based amplification. All these methodological aspects make the data of this study very robust.

Our analysis was triggered by the simple observation that the IgM response to SARS-CoV-2 antigens in our cohort did not precede the IgA and IgG responses as expected. In fact, in the first 2020 reports of antibody response to SARS-CoV-2, this anomaly was already detected, but the focus was on applying serology to diagnosis. ^9,10,44,45^

A close analysis of the antibody response trajectories by SARS-CoV2 protein and isotype, stratified by patients’ severity, offers additional cues of a recall immune response. The fast and dominant IgA responses to the Mpro, NP, and Spike suggest a mucosal origin from memory resident T (Trm) and B cells in the mucosa of the upper airways.^46–48^ That the IgM response, even if weaker, is dominated by the response to Spike, whose sequence is more different from that of CCCV, suggests the recruitment of naïve cells, even if late, contributes to mounting a response to SARS-CoV-2 specific epitopes. The vigorous but late IgG response may originate from deeper lymph nodes draining the lower respiratory airways. The response to RBD is challenging to analyse due to the much smaller antigen size used in the assay. It is, however, noteworthy that we detected a robust late IgG response to RBD in moderate and severe patients. Another, not mutually exclusive, possibility is that SARS-CoV-2 Spike can engage naïve lymphocytes in the mucosa where tissue damage generates more PAMP and jump-starts the immune response rather than originating from Trm.^47^ All the above supports the concept that in COVID-19, a recall response to antigens cross-reactive with CCCV coexists with a response to new epitopes in the SARS-CoV-2 Spike protein.

The limitation of our study is that we did not measure antibodies to endemic coronavirus in samples obtained before and after the COVID-19 episode as required to demonstrate immune imprinting. However, a contemporary study in the same populations showed a high prevalence of antibodies to endemic coronavirus in the same population.^17^ Another limitation is that we did not test the sera for neutralising SARS-CoV-2 antibodies, but it has been repeatedly shown that they correlate closely with anti-RBD antibodies. ^49^

Antigenic original sin, as the negative aspect of IP, may be deleterious to the host. The predominant immune response is directed to the dominant epitopes of the prior immunising virus, and the effectors generated, antibodies and CD8 T cells, having low affinity for the new epitopes, are inefficient, resulting in a more severe infection. The conceptual cellular and molecular basis is well understood; memory B and T cells capture/recognise the cross-reactive protein/peptide, and as they have a lower threshold for activation, they dominate the response, preventing the activation of naïve cells that may carry more specific receptors for the new epitopes. However, because medium affinity antibodies and T cells may have some protective effect, IP sin does not determine a severe infection. ^50^ In addition, as the amount of viral antigens reaching the lymph nodes increases with time, a primary specific response eventually emerges, takes over, and leads to the control of the infection. Besides interfering with the primary response, IP can be associated with antibody-dependent enhancement (ADE). Observed in viral infections, e.g., dengue, and occasionally after vaccination, ^51^ ADE is attributed to macrophage infection via Fc and complement receptors, leading to infection dissemination and M2 polarisation.^52^ The participation of ADE in COVID-19 has been discussed, and it has been a concern for developing the SARS-CoV-2 vaccines. ^53^ Additionally, original antigenic sin, generating low-affinity antibodies to coronavirus proteins, may also favour immunocomplex formation and coagulation dysregulation. ^54^

The observations from our study and others ^13,16,17,21,43,53,55–58^ that suggest an element of IP / original antigenic sin in COVID-19 can be summarised as follows:

1. The lack of preceding IgM responses in most cases.
2. The quick fall and short life of antibody response, a feature of IP. ^18^
3. The response to RBD is particularly delayed and subject to large interindividual variability.
4. The correlation of SARS-CoV-2 titres with disease severity and not with viral load^59^ supports the contention that in many cases the antibody response contributes to severity rather than resolution of the infection; this would fit with a response directed in part to epitopes cross-reactive with CCCV.
5. Plasmablast levels increase significantly in the more severe categories, reminiscent of dengue at second exposure. ^60^
6. The preferential expansion of surface-negative plasmablasts suggests that memory B cells, not naive cells, in the mucosa and deep SLO are mobilised as pre-plasma cells. ^61^
7. The features of B cell subsets and cTfh profiles and their correlation with the antibody responses are more consistent with a recall than a primary response.

In conclusion, upon analysing the trajectory of a set of immunological variables during the acute episode of COVID-19, we can infer that in most cases, the immune response to SARS-CoV-2 is a recall response previous exposure to endemic coronavirus. This opens the possibility of reinterpreting the immunopathology of COVID-19 supporting a more important role of IP (McNaughton 2022,Rijkers 2021,Tetro 2020) as determinant of severity.

## AUTHOR CONTRIBUTION

D. A-S, M. M-G, A. S-M, J. B-M, A. T-S, M. H-G, and R. P-B conceived and designed the project. A. S-M designed the redcap database. M.A. F-S and H. A performed the high-dimension flow cytometry data analysis and interpretation. J.A.E-P., E.P-C. C. Z-E, F.M-R, and B. U-V collected samples and clinical data from hospitalised patients, while C.V. organised it from the primary care centres and visiting teams. R.P. organised the control group. A. S-P. conducted the statistical and bioinformatic analysis. E.M-C., M. M-G, and C.V. reviewed and contributed to the manuscript drafts. P.K. reviewed the pre-final versions and suggested insightful analyses. A.V. conducted the Nanostring analysis n. D. A-S ran the project’s logistics, conducted the cytokine measurements, and performed some statistical and bioinformatic analyses. R.P-B supervised the project, carried out the global data analysis and interpretation, wrote the manuscript, and prepared tables and figures.

## Data Availability

All data produced in the present study are available upon reasonable request to the authors

## ACKNOWLEDGMENTS

This study was funded by grants COV20/00416, Cov20/00654, and COV20/00388 to RP-B, AT-S, and JB-M respectively, from Instituto de Salud Carlos III (ISCIII), Ministry of Economy and Competitiveness, Spain, co-financed by the European Regional Development Fund (ERDF). D.Á-S was the recipient of a doctoral fellowship from the Vall d’Hebron Research Institute (VHIR), Barcelona, Spain, up to 2023 and from 2024 by a ISCIII postdoctoral fellowship, “Sara Borrell,” CD23/0011. A.S-M was supported by a ISCIII postdoctoral fellowship, “Juan Rodés” (JR18/00022). Bioinformatics analysis has been carried out in part in the Statistics and Bioinformatics Unit (UEB) at Vall d’Hebron Research Institute (VHIR).

The authors thank all the patients and health staff of the Hospital Universitari Vall d’Hebron, Hospital Universitari Bellvitge, and Hospital Universitari Germans Trias i Pujol and Primary Care Units who endured the pandemic and did their best to overcome the successive waves of COVID-19 in Barcelona, an experience that none of us would ever forget. The authors are grateful to Sergio Navarro Velázquez and Mario Framil Seoane from Hospital Universitari Bellvitge and to Arturo Llobell Uriel and Joan Roig Sanchis from Hospital Universitari Vall d’Hebron for their support in reviewing patients’ records within the general Immuno-profile COVID-ICS project. Dr Isabel Novoa-Garcia and Ms. Sheyla Pascual Martin for their invaluable help in organising and maintaining the COVID-19 collection in the HUVH BioBank. To Ms Adelaida Parada and the administrative and technical staff of immunology laboratories at the participating hospitals that collected and organised the COVID-19 patient samples for review by the medical personnel of this study. Artificial Intelligence-based software Grammarly® has only been used to improve the readability of some paragraphs by R.P-B; the text was reviewed, and the authors take full responsibility for the final version.

## DATA SHARING

Supplemental tables contain the additional data required to re-analyse the data: the xlsx file with the transcriptomic data will be made available on request.

## Supplemental figures and legends

**Supplemental Figure S1.**
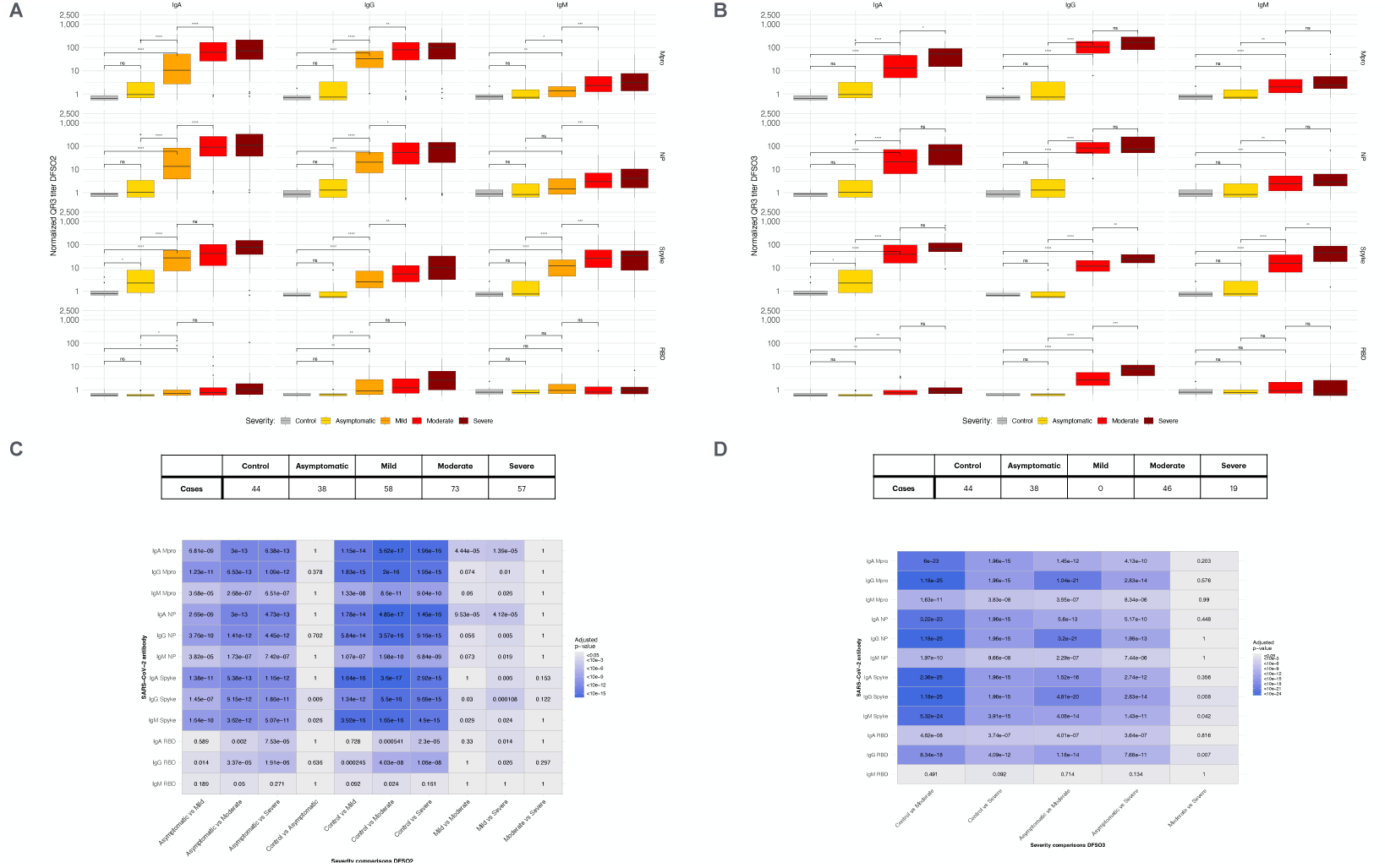
Pairwise comparison of antibody response to SARS-CoV-2 proteins in the different groups of patients. (**A)** and **(B**) Plots corresponding to the 8-20 days period (DFSO2) from symptom onset and 0-107 days (DFSO3). Median ± IQR. (**C**) and (**D**) Tables with the number of cases in each category and adjusted *p* values for each possible comparison from (**A**) and (**B**). Pairwise Wilcoxon rank sum test. The corresponding graphics for period DFSO1 are in Figure 1.

**Supplemental Figure 2.**
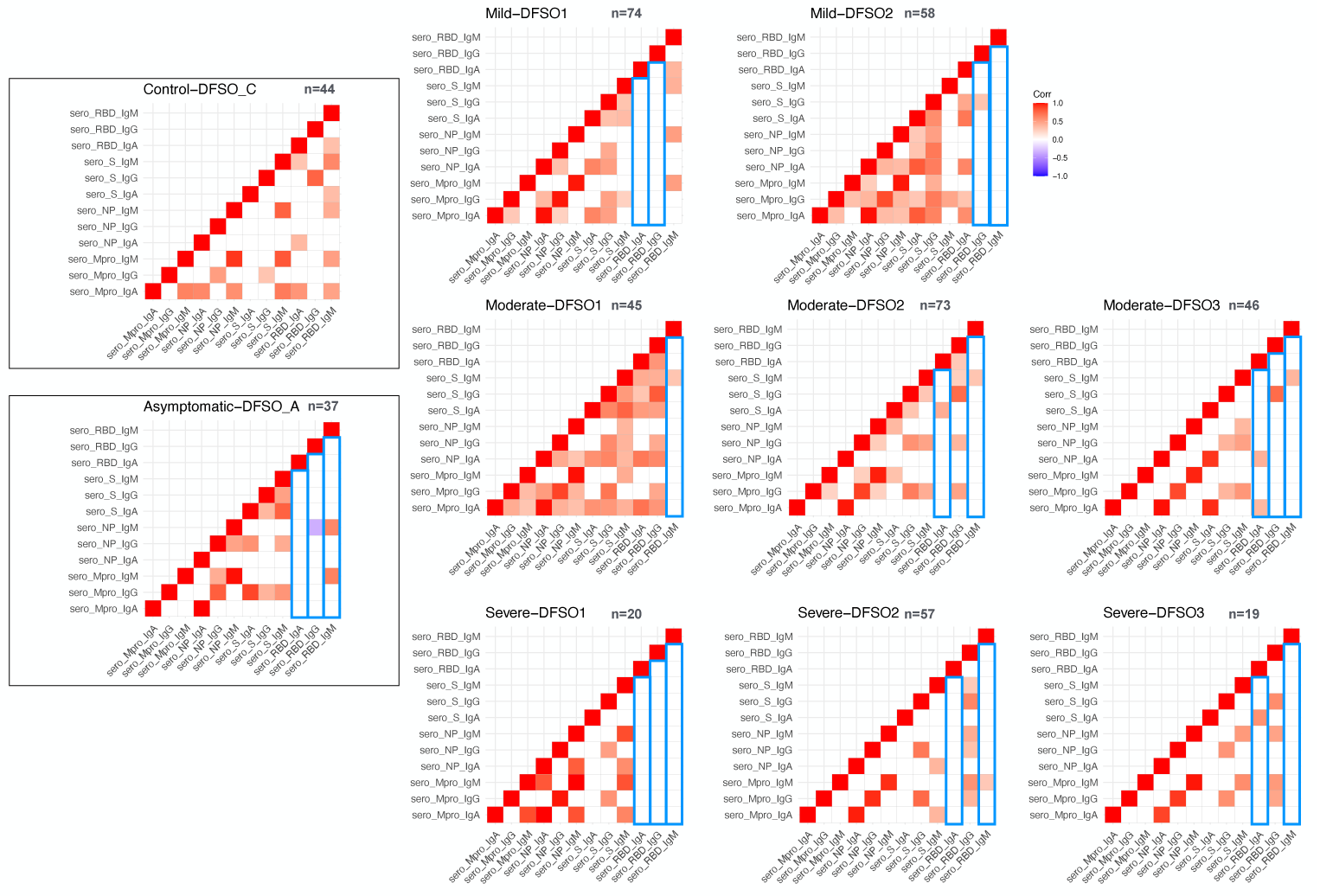
Mutual correlation among the antibody responses to SARS-CoV-2 by antigen and Ig isotype stratified by severity and time (Spearman). The ten panels show mutual correlations; the number of patients is indicated in the upper right corner of each panel. DFSO, days from symptoms onset; DFSO1: 0-7, DFSO2:8-20, DFSO3: 21-107. In addition, DFSO_C corresponds to controls, and DFSO_A corresponds to asymptomatic. Maximal mutual correlations are seen in the mild and moderate panels, and correlations increase in severe patients at the late time point. RBD is mostly dissociated, highlighted by the blue boxes.

**Supplemental Figure 3:**
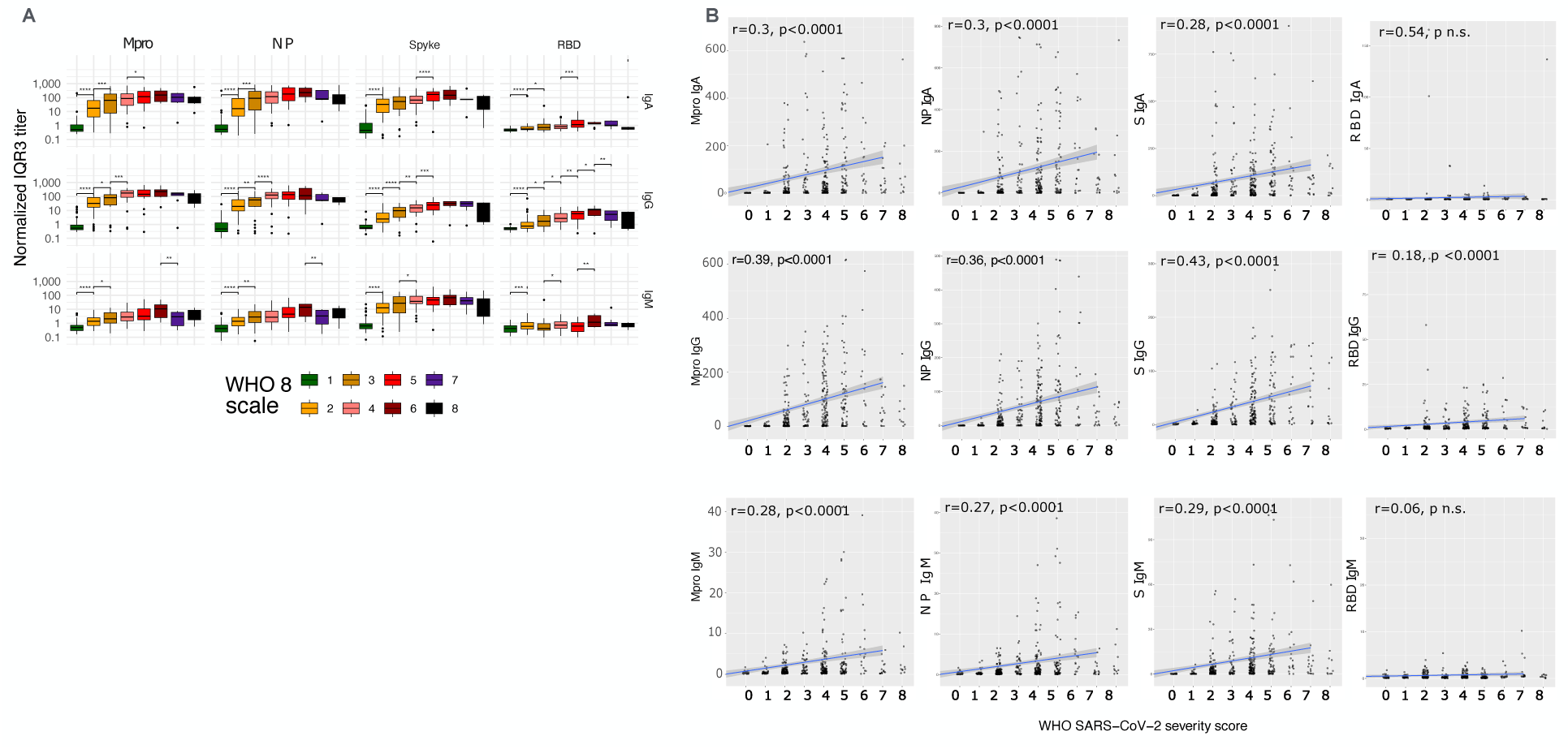
SARS-CoV-2 antibody titers and WHO severity scale. (**A**) Only significant differences to the adjacent groups are annotated in box plot severity group comparisons. Median ± IQR. (B) Panels of linear correlation analysis for each antibody to SARS-CoV-2 proteins. X-axis WHO classification has been used as a continuous variable for this analysis. Y-axis normalized antibody titers. Notice that all correlations are positive, but only anti-S IgG severity shows an r>0.4; the correlation does not extend beyond category 6; critically ill patients had lower antibody levels.

**Supplemental Figure 4:**
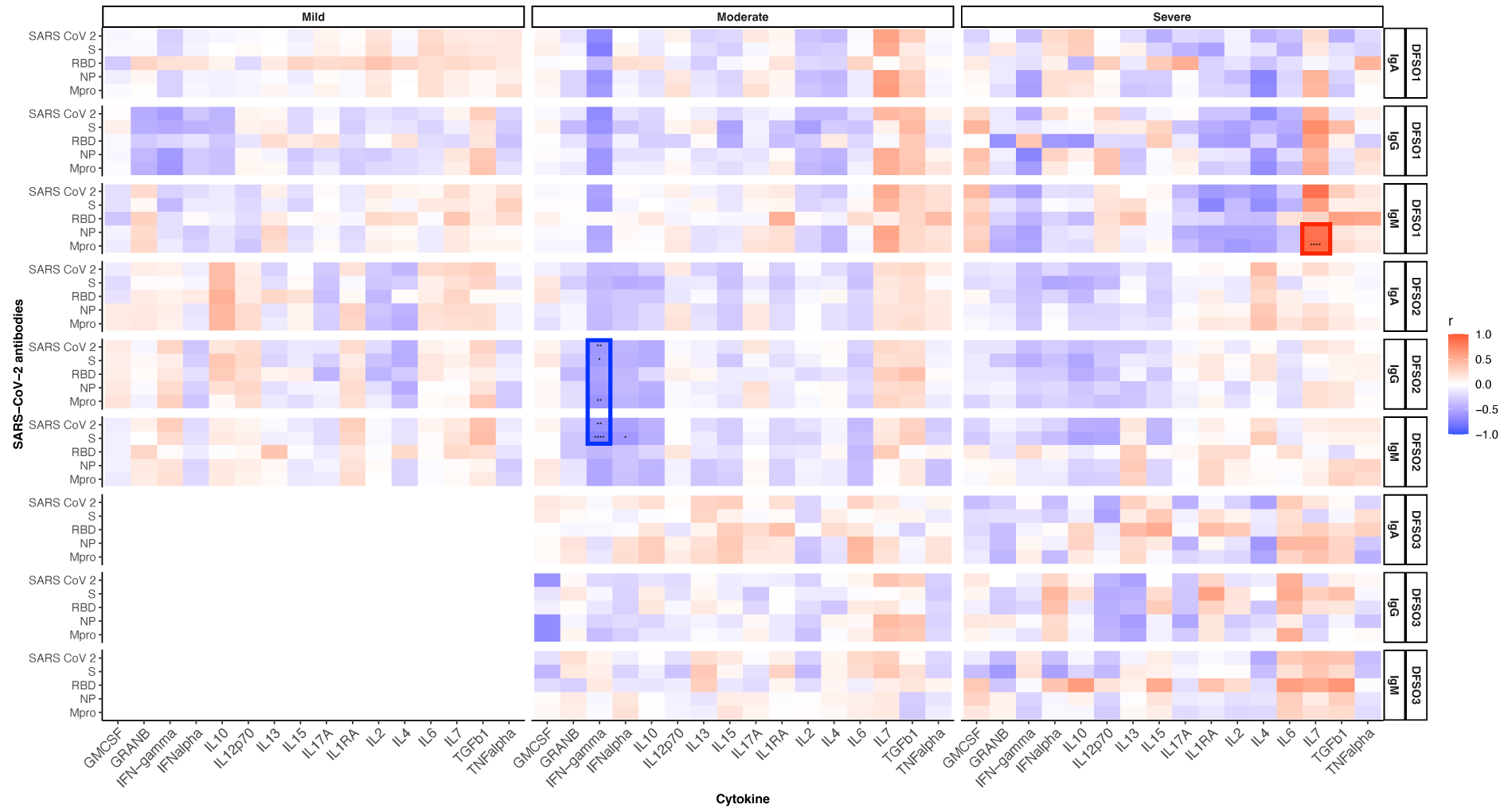
Correlation among the antibody responses to SARS-CoV-2 by antigen, Ig isotype, and cytokines stratified by severity and time. Data from 309 samples corresponding to 159 representative cohort cases were available. There is a negative correlation of IFNs, especially gamma, with the serological response (blue box). Correlation of IL-7 to serological response (red box), see text for the interpretation.

**Supplemental Figure 5.**
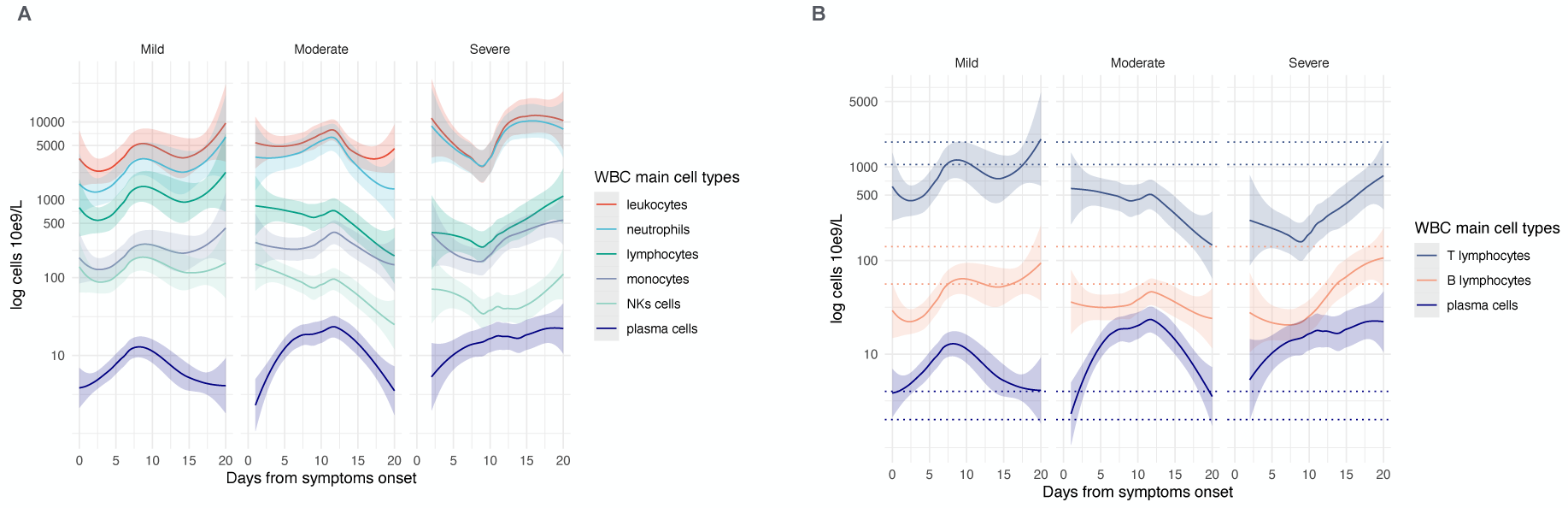
LOESS trajectories of the main differential white blood cell count by severity groups during the initial 20 days. The lines represent values averaged by LOESS regression of 326 samples collected before day 20; the number of samples by severity group was 133 Mild, 155 Moderate, and 78 Severe. The CI has been adjusted to 0.75 to reduce overlaps. The y-axis is on a log scale, but the labels correspond to the number of cells; values from controls IQR 0.25 – 0.75 are indicated by horizontal dots or dash lines that maintain the color code. In (A), the four main types and plasmablast/plasma cells are plotted; three populations are plotted in (B) to compare the trajectories.

**Supplemental Figure 6.**
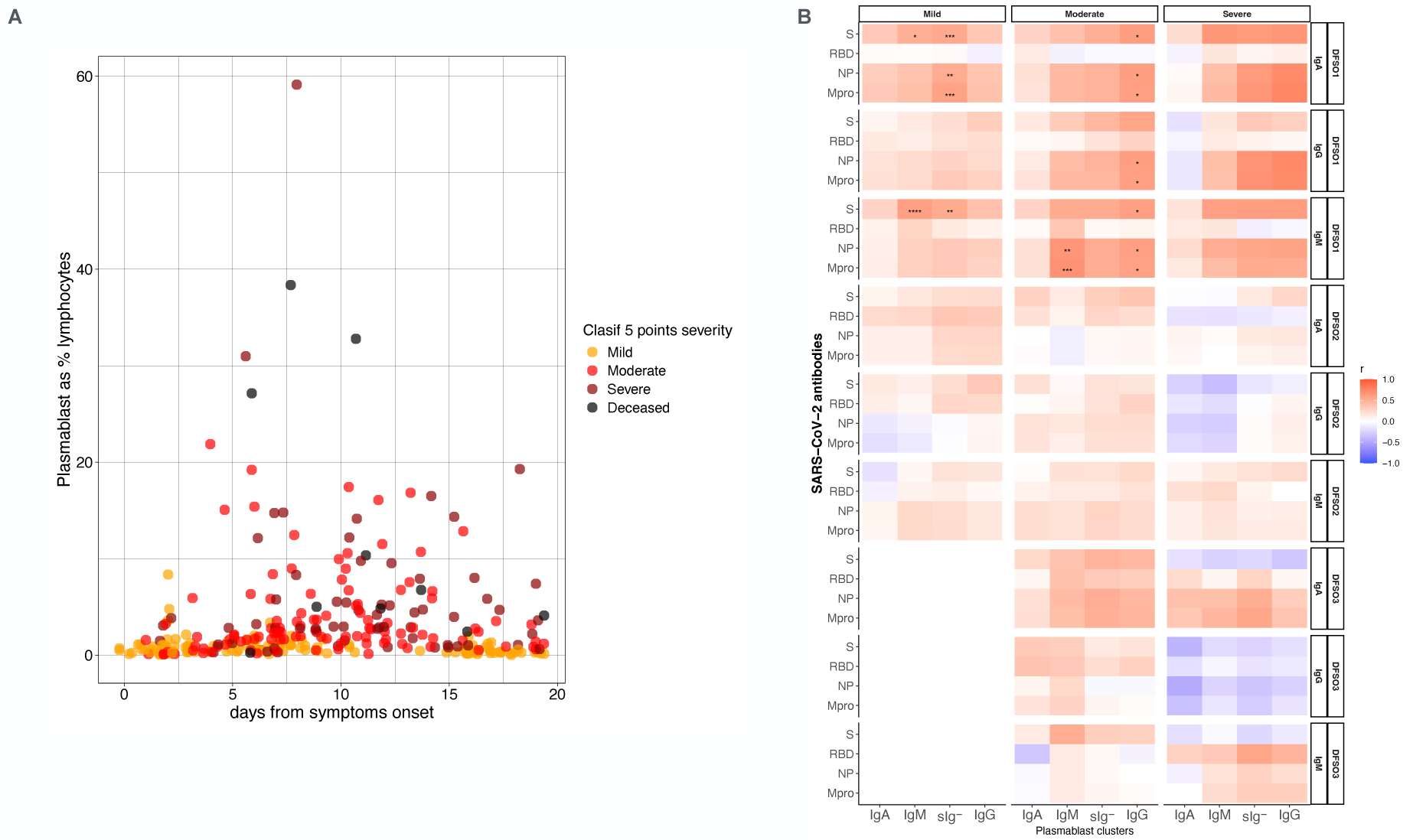
Plasmablast (PB). (**A**) Dot plot showing the percentage of total PBs in the cohort of symptomatic COVID-19 patients during the initial 20 days of follow-up. Extreme values of up to 60% of the lymphocytes are observed in severe and deceased extremely lymphopenic patients. (B) Distribution of subsets in PB from controls in which total PB makes only 0.16 [0.09 - 0.27] % of the lymphocytes.

**Supplemental Figure 7.**
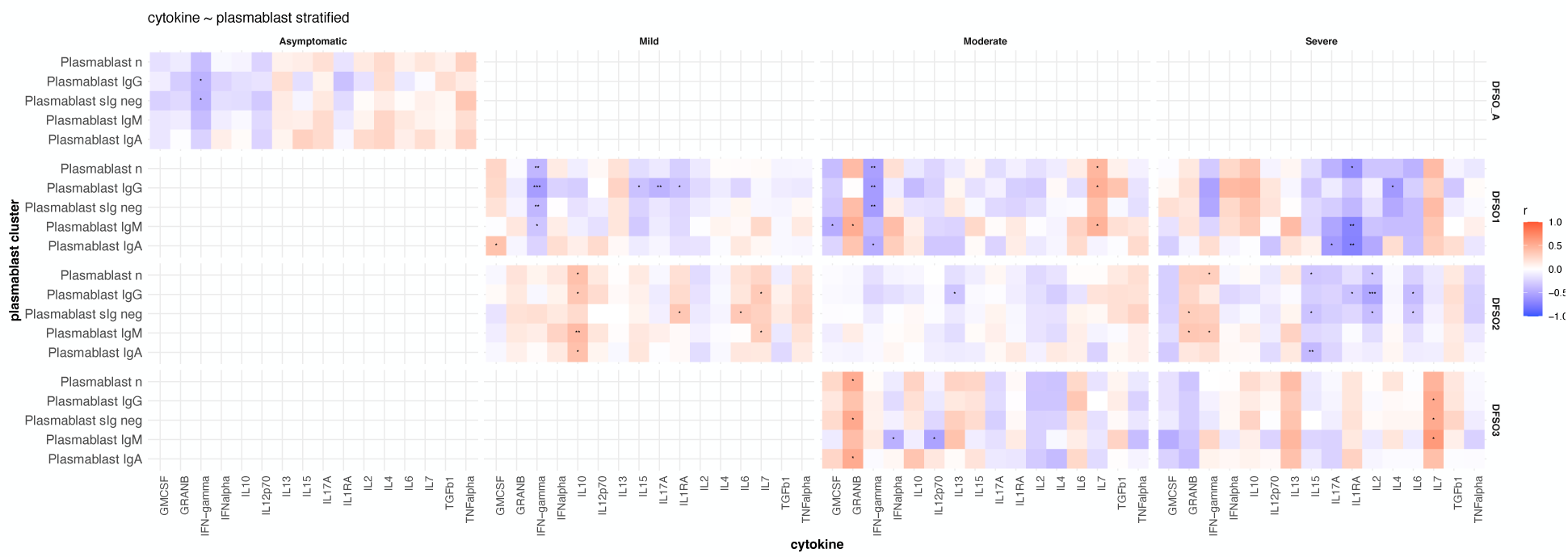
Correlation of cytokines with plasmablast subsets. DFSO, Days from symptom onset. DFSO1, samples collected between 0-7 days; DFSO2, samples collected between 8-28 days; DFSO3, samples collected between 28-107 days. See text for comments. Adjusted *p* values p <0.05=*, *p* < 0.01=**, *p* < 0.001=***, *p* < 0.0001****, p not adjusted. See comments in the text.

**Supplemental Figure 8.**
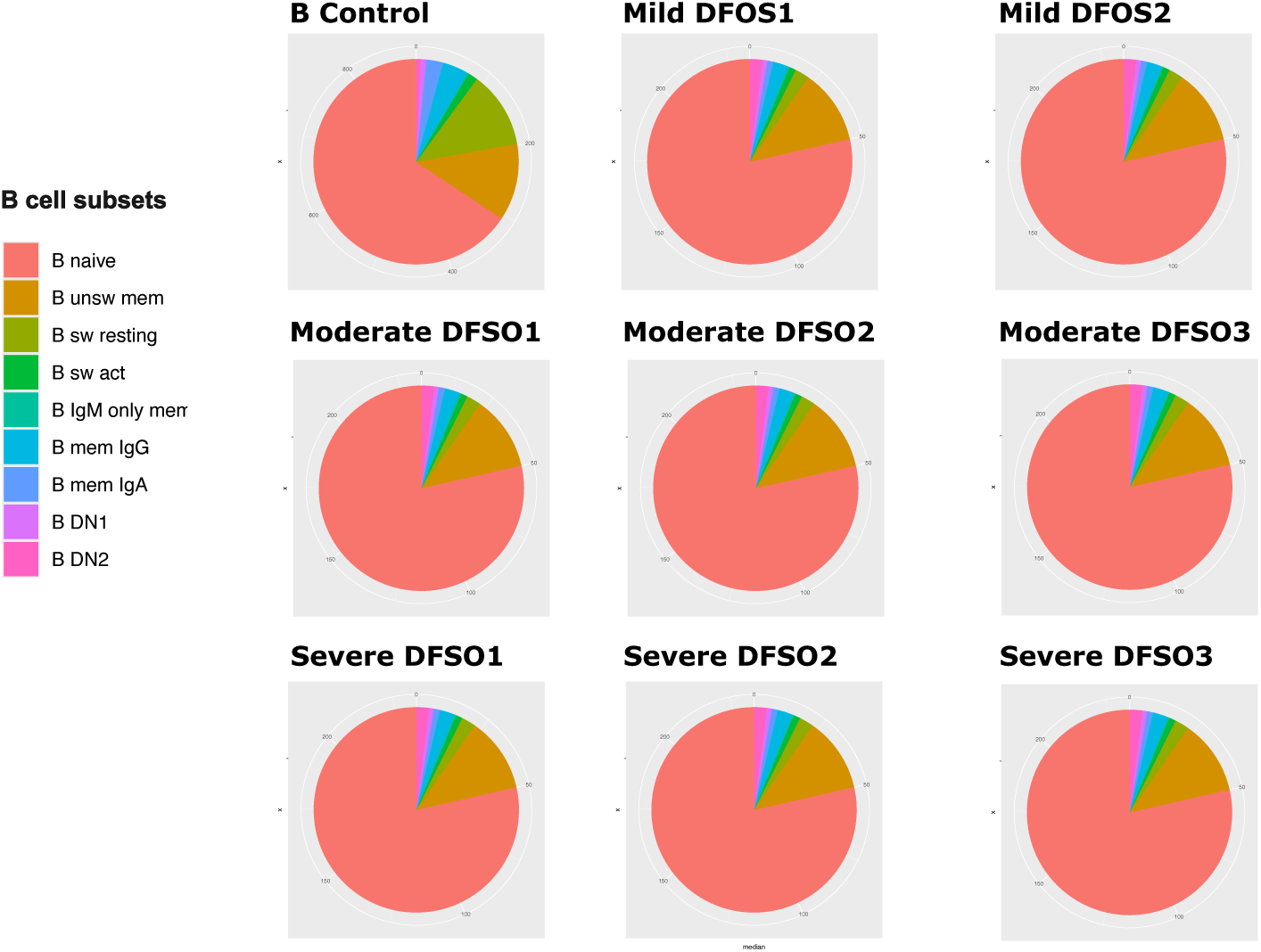
Distribution of B cell subsets in controls and patients. A set of pie plots show the changes in the distribution of B cells among the different subsets during the COVID-19 time-course. Notice that B cell switched resting is the only population showing a major reduction. Abbreviations: DFSO, Days from symptoms onset; unsw, unswitched; sw, switched; act, activated; mem, memory, DN, doble negative (IgD^-^CD27^-^).

**Supplemental Figure 9.**
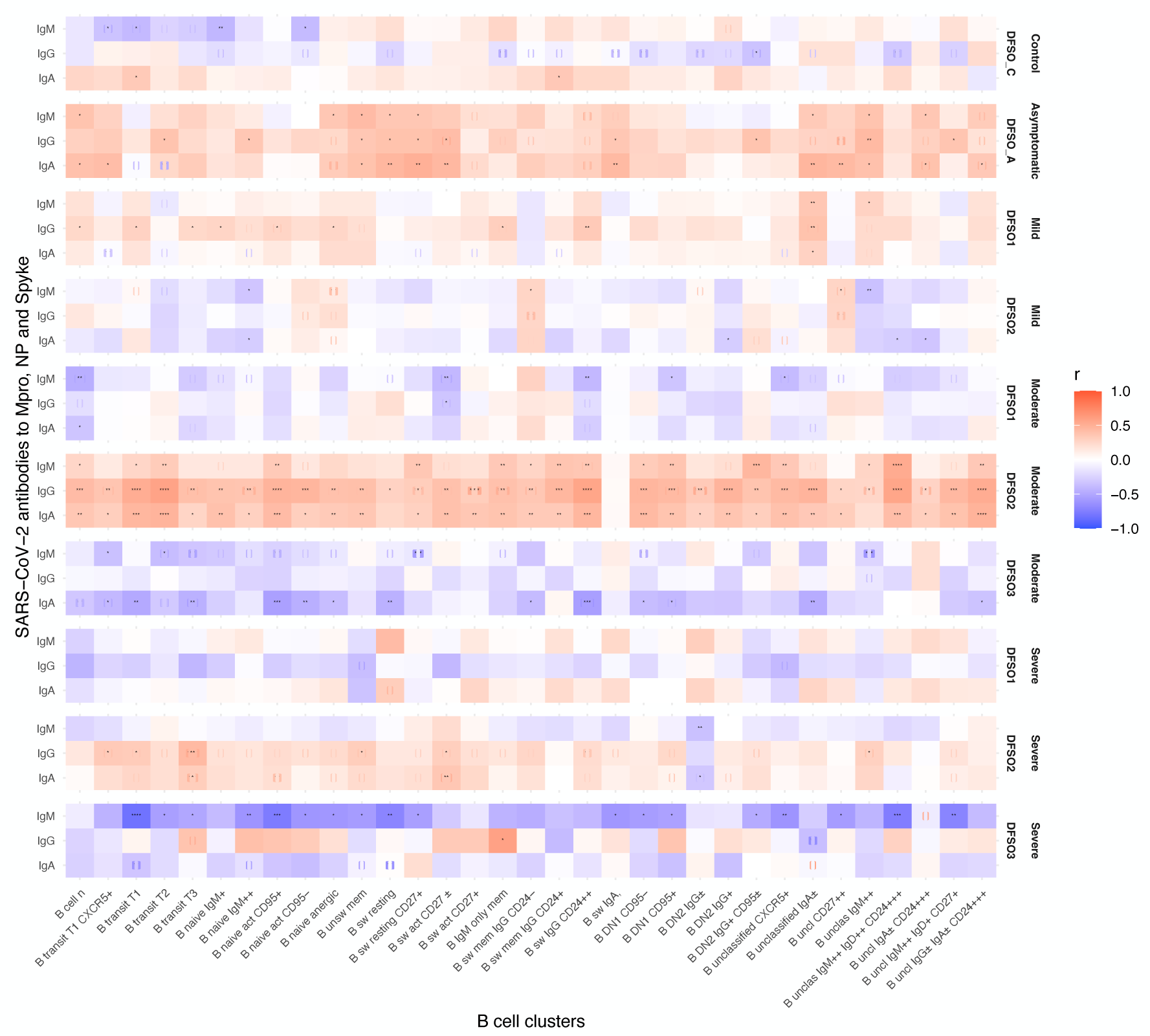
Correlation of B cell cluster and serological response. DFSO_C and DFSO_A period during which control and asymptomatic patients’ samples were collected; DFSO1, samples collected between 0-7 days; DFSO2, samples collected between 8-28 days; DFSO3, samples collected between 28-107 days. See text for comments. Adjusted *p*, *p*<0.05 = *, *p* <0.01 = **, *p*<0.001 = ***, *p*< 0.0001 = ****. See comments in the text.

**Supplemental Figure 10.**
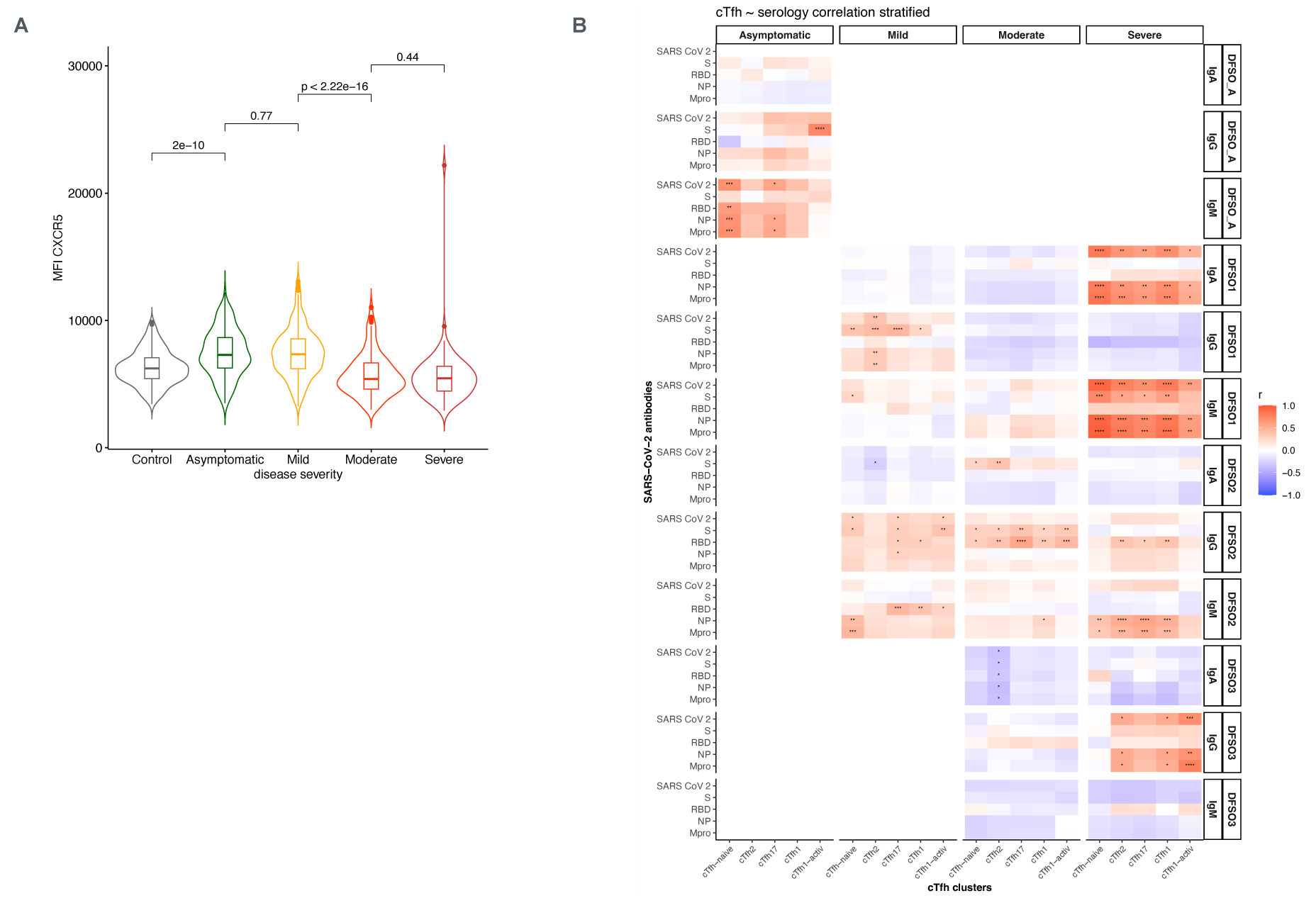
cTfh and serological response (Spearman). (**A**) CXCR5 expression level (MFI) of cTfh in the different severity groups. Pairwise Wilcox test. Median ± IQR. (B) Correlation of cTfh cell number and serological responses by severity and time period. ****. Notice that significant correlations are clustered for IgM in asymptomatic and for IgM, IgA, and IgM in the different periods of severe patient’s follow-up. See text for comments. *p* < 0.05= *, *p* < 0.01 =**, *p* < 0.001=***, *p* < 0.0001 = ****.

**Supplemental Figure 11.**
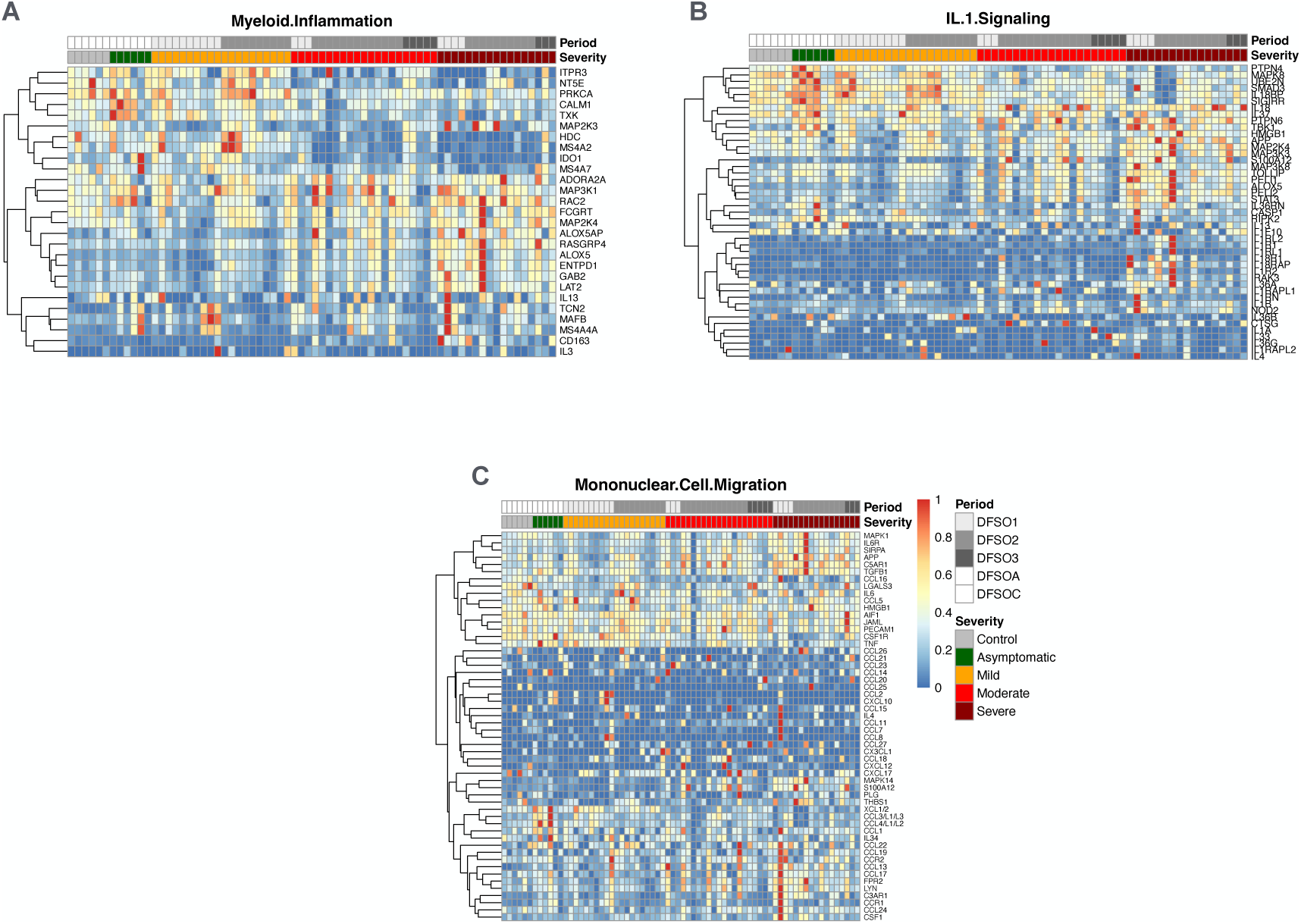
Heatmaps summarizing the transcriptomic profile of 33 COVID-19 patients stratified by severity and follow-up periods. (A) Myeloid inflammation; (B) IL-1 signaling; and (C) Mononuclear cell Migration.

---

**Supplemental Table 1.**
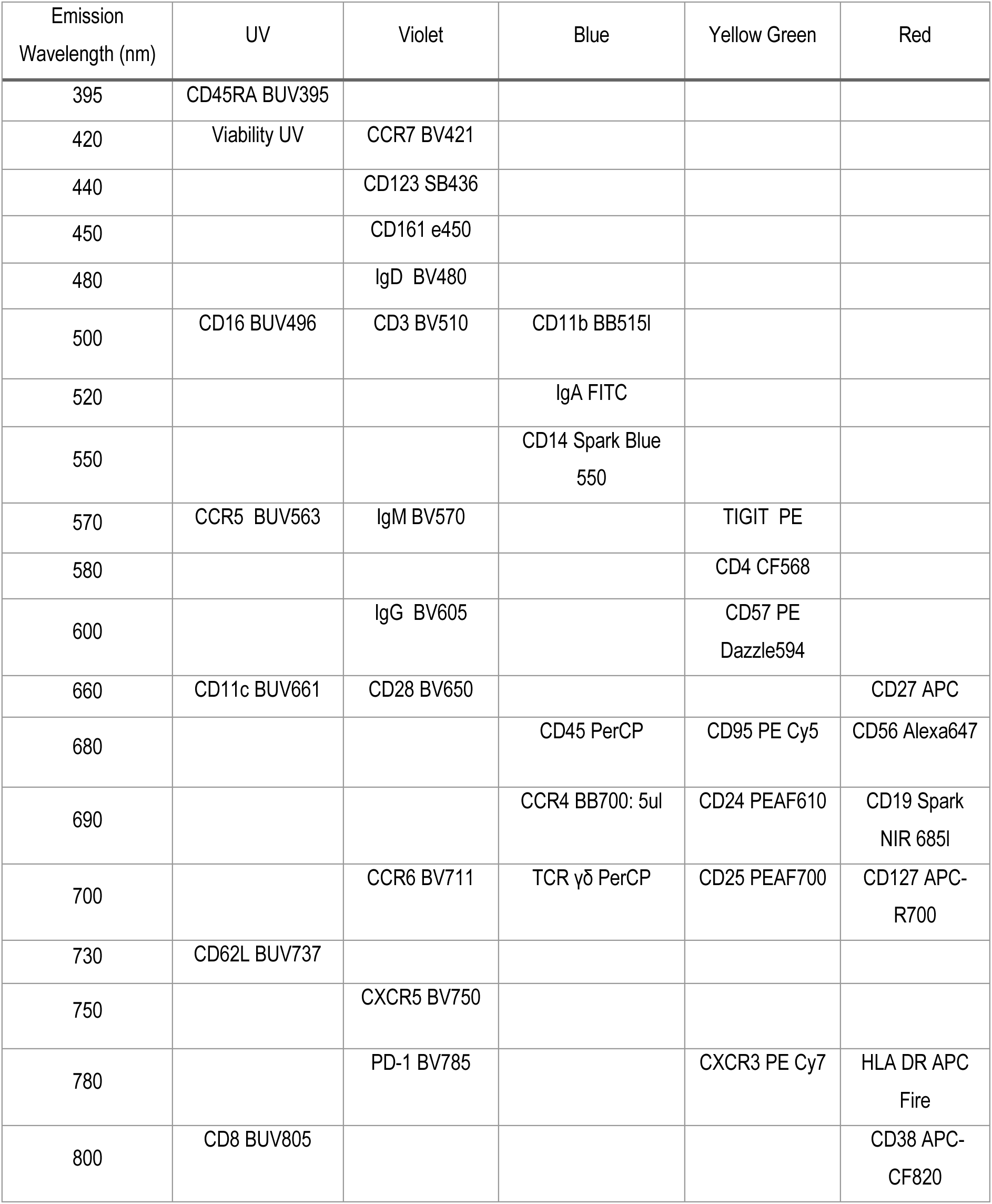
Panel Flow Cytometry Reagents.

**Supplemental Table 2.** B cell subset statistics.

| Period | Subset B cell | Compared groups |  | p.adj |
| --- | --- | --- | --- | --- |
| DFS01 | B cell n | Mild | Moderate | 1.16E-07 |
| DFS01 | B cell n | Mild | Severe | 5.61E-21 |
| DFS01 | B cell n | Moderate | Severe | 1.31E-08 |
| DFS02 | B cell n | Mild | Moderate | 1.58E-33 |
| DFS02 | B cell n | Mild | Severe | 6.99E-08 |
| DFS02 | B cell n | Moderate | Severe | 1.00E-03 |
| DFS03 | B cell n | Moderate | Severe | 4.00E-03 |
| DFS01 | B transitional | Mild | Moderate | 9.00E-03 |
| DFS01 | B transitional | Mild | Severe | 1.08E-79 |
| DFS01 | B transitional | Moderate | Severe | 1.07E-51 |
| DFS02 | B transitional | Mild | Moderate | 2.51E-56 |
| DFS02 | B transitional | Mild | Severe | 5.70E-57 |
| DFS02 | B transitional | Moderate | Severe | 2.55E-01 |
| DFS03 | B transitional | Moderate | Severe | 2.38E-65 |
| DFS01 | B naive | Mild | Moderate | 1.00E+00 |
| DFS01 | B naive | Mild | Severe | 2.00E-17 |
| DFS01 | B naive | Moderate | Severe | 6.72E-15 |
| DFS02 | B naive | Mild | Moderate | 2.28E-41 |
| DFS02 | B naive | Mild | Severe | 1.14E-05 |
| DFS02 | B naive | Moderate | Severe | 9.33E-14 |
| DFS03 | B naive | Moderate | Severe | 1.20E-01 |
| DFS01 | B unswitched mem | Mild | Moderate | 1.37E-13 |
| DFS01 | B unswitched mem | Mild | Severe | 4.62E-11 |
| DFS01 | B unswitched mem | Moderate | Severe | 1.00E-02 |
| DFS02 | B unswitched mem | Mild | Moderate | 8.49E-21 |
| DFS02 | B unswitched mem | Mild | Severe | 1.70E-02 |
| DFS02 | B unswitched mem | Moderate | Severe | 2.30E-05 |
| DFS03 | B unswitched mem | Moderate | Severe | 1.63E-01 |
| DFS01 | B IgM only mem | Mild | Moderate | 3.15E-70 |
| DFS01 | B IgM only mem | Mild | Severe | 6.78E-25 |
| DFS01 | B IgM only mem | Moderate | Severe | 6.00E-03 |
| DFS02 | B IgM only mem | Mild | Moderate | 9.99E-84 |
| DFS02 | B IgM only mem | Mild | Severe | 1.78E-63 |
| DFS02 | B IgM only mem | Moderate | Severe | 1.00E+00 |
| DFS03 | B IgM only mem | Moderate | Severe | 2.40E-01 |
| DFS01 | B mem IgA | Mild | Moderate | 2.02E-106 |
| DFS01 | B mem IgA | Mild | Severe | 5.88E-39 |
| DFS01 | B mem IgA | Moderate | Severe | 2.65E-01 |
| DFS02 | B mem IgA | Mild | Moderate | 8.58E-72 |
| DFS02 | B mem IgA | Mild | Severe | 5.88E-79 |
| DFS02 | B mem IgA | Moderate | Severe | 7.89E-01 |
| DFS03 | B mem IgA | Moderate | Severe | 3.96E-05 |
| DFS01 | B mem IgG | Mild | Moderate | 1.85E-72 |
| DFS01 | B mem IgG | Mild | Severe | 6.09E-60 |
| DFS01 | B mem IgG | Moderate | Severe | 1.10E-07 |
| DFS02 | B mem IgG | Mild | Moderate | 7.05E-119 |
| DFS02 | B mem IgG | Mild | Severe | 7.50E-123 |
| DFS02 | B mem IgG | Moderate | Severe | 2.00E-03 |
| DFS03 | B mem IgG | Moderate | Severe | 1.07E-01 |
| DFS01 | B switched resting | Mild | Moderate | 3.36E-79 |
| DFS01 | B switched resting | Mild | Severe | 1.48E-63 |
| DFS01 | B switched resting | Moderate | Severe | 1.75E-05 |
| DFS02 | B switched resting | Mild | Moderate | 2.43E-85 |
| DFS02 | B switched resting | Mild | Severe | 7.14E-69 |
| DFS02 | B switched resting | Moderate | Severe | 1.00E+00 |
| DFS03 | B switched resting | Moderate | Severe | 3.69E-05 |
| DFS01 | B switched act | Mild | Moderate | 4.83E-114 |
| DFS01 | B switched act | Mild | Severe | 1.04E-45 |
| DFS01 | B switched act | Moderate | Severe | 4.20E-01 |
| DFS02 | B switched act | Mild | Moderate | 8.16E-73 |
| DFS02 | B switched act | Mild | Severe | 3.84E-84 |
| DFS02 | B switched act | Moderate | Severe | 7.10E-02 |
| DFS03 | B switched act | Moderate | Severe | 6.04E-01 |
| DFS01 | B DN1 | Mild | Moderate | 4.92E-15 |
| DFS01 | B DN1 | Mild | Severe | 2.44E-28 |
| DFS01 | B DN1 | Moderate | Severe | 1.93E-08 |
| DFS02 | B DN1 | Mild | Moderate | 9.03E-39 |
| DFS02 | B DN1 | Mild | Severe | 3.06E-46 |
| DFS02 | B DN1 | Moderate | Severe | 4.10E-02 |
| DFS03 | B DN1 | Moderate | Severe | 9.40E-05 |
| DFS01 | B DN2 | Mild | Moderate | 6.78E-17 |
| DFS01 | B DN2 | Mild | Severe | 7.77E-01 |
| DFS01 | B DN2 | Moderate | Severe | 8.13E-06 |
| DFS02 | B DN2 | Mild | Moderate | 3.33E-17 |
| DFS02 | B DN2 | Mild | Severe | 6.00E-02 |
| DFS02 | B DN2 | Moderate | Severe | 3.93E-21 |
| DFS03 | B DN2 | Moderate | Severe | 4.05E-19 |
| DFS01 | B unclassified | Mild | Moderate | 1.81E-17 |
| DFS01 | B unclassified | Mild | Severe | 3.99E-13 |
| DFS01 | B unclassified | Moderate | Severe | 1.52E-01 |
| DFS02 | B unclassified | Mild | Moderate | 4.74E-38 |
| DFS02 | B unclassified | Mild | Severe | 3.75E-06 |
| DFS02 | B unclassified | Moderate | Severe | 1.36E-10 |
| DFS03 | B unclassified | Moderate | Severe | 1.59E-04 |

